# Enhancing COVID-19 Forecasting Precision through the Integration of Compartmental Models, Machine Learning and Variants

**DOI:** 10.1101/2024.03.20.24304583

**Authors:** Daniele Baccega, Paolo Castagno, Antonio Fernández Anta, Matteo Sereno

## Abstract

Predicting epidemic evolution is essential for informed decision-making and guiding the implementation of necessary countermeasures. Computational models are vital tools that provide insights into illness progression and enable early detection, proactive intervention, and targeted preventive measures.

This paper introduces Sybil, a framework that integrates machine learning and variant-aware compartmental models, leveraging a fusion of data-centric and analytic methodologies. To validate and evaluate Sybil’s forecasts, we employed COVID-19 data from two European countries. The dataset included the number of new and recovered cases, fatalities, and variant presence over time. We evaluate the forecasting precision of Sybil in periods in which there is a change in the trend of the pandemic evolution or a new variant appears. Results demonstrate that Sybil outperforms a conventional data-centric approach, being able to forecast accurately the changes in the trend, the magnitude of these changes, and the future prevalence of new variants.

## Introduction

The COVID-19 pandemic, caused by the SARS-CoV-2 virus, highlights the intricate challenges of addressing the most impactful global health crisis of the 21st century. The rapid global spread of the virus has affected nearly every part of the world. Consequently, healthcare systems worldwide are grappling with the significant challenge posed by COVID-19, requiring a continuous COVID-19 monitoring system that includes robust surveillance, widespread testing, contact tracing, and can be used to plan and deploy stringent infection control measures.

Establishing a continuous monitoring system aids policymakers in effectively managing the socio-health emergency brought about by the epidemic. Accurate forecasting is a fundamental element of such a system, and is crucial for efficient planning, resource allocation, and decision-making within public health authorities. It facilitates the development of proactive measures, such as vaccination campaigns, travel advisories, and community engagement programs, fostering public awareness and participation in disease control efforts. This proactive approach enhances preparedness and is critical in curbing the spread of infectious diseases and mitigating their impact on communities worldwide. Epidemic forecasting involves a multidisciplinary approach, integrating epidemiology, mathematical modeling, data analysis, and computational methods to gain insights into the future progression of outbreaks.

Numerous methodologies are available for predicting the future trajectory of an epidemic, leveraging diverse modeling approaches. Particularly, machine learning (ML) models^1–3^ and especially deep learning (DL) models, including Convolutional **Neural Networks (CNNs), Recurrent Neural Networks (RNNs) featuring Long Short-Term Memory (LSTM) or Gated Recurrent** Unit (GRU) cells, and multivariate CNNs^4–6^, have emerged as highly prominent approaches for forecasting. Several studies use a combination of multiple data-centric approaches (e.g., a machine learning model with ARIMA or Prophet^7^). Despite their increasing popularity, surpassing conventional techniques such as auto-regressive models (e.g., ARIMA, SARIMA)^2,5,8^, these data-centric approaches exhibit notable limitations. First of all, predictions are not easily explainable, and secondly, they often fail to predict changes in the trend, such as peaks or the appearance of new variants.

On the other hand, compartmental models^9–11^ and stochastic transmission models^12^ are analytic approaches specifically tailored to reproduce the evolution of an infection in a population and in the presence of variants^13–16^. These models incorporate factors such as population demographics, rates of infection, recovery, and mortality, providing a mathematical representation of epidemic progression^17,18^. This transparency simplifies explaining and interpreting the results to policymakers, healthcare professionals, and the general public, thus fostering a better understanding and informed decision-making. Indeed, several COVID-19 forecasting studies incorporate data-centric and analytical approaches, such as a combination of compartmental models and ARIMA^19^, and several other combinations^20^.

Forecasting the epidemic spread aims to accurately predict the percentage of the infected population at a given point in the future, the number of fatalities, hospitalizations, and so on. However, these percentages result from complex population dynamics that often show nonlinear behaviors, particularly at critical points where there are changes in the infection course— such as at the peak of the diffusion or when a new variant arises. Conversely, epidemics dynamics are often characterized by widely recognized quantities. The most well-known is the basic reproduction number, R_0_, which express the number of secondary infection arising from one single infected individual within a population of susceptible individuals. Although R_0_ is useful to characterize if and how fast a disease spreads in a population, its time-dependent counterpart, R_t_, enables a quantitative evaluation of the infection course. Such indicators, being specific to the disease, tend to be stable. Therefore, their future evolution shows a more predictable behavior.

Following the approach of combining data-driven approaches with analytical models, we propose **Sybil**, an integrated machine learning and variant-aware compartmental model framework capable of providing improved prediction accuracy and explainability. Sybil exploits the relative stability of disease characteristics indices to project in the future and employs a simple and widely recognized analytical model to draw the infection dynamic. Sybil’s strengths mark the difference with approaches present in the literature thanks to *i)* its capability of providing accurate forecasts, even when there are relevant changes in the diffusion process, and *ii)* reduced need for training data. Furthermore, the approach offers *iii)* the possibility to study the evolution of the infection of several variants and *iv)* the replicability of the results. Additionally, *v)* the open-source code is available online.

The joint use of ML and analytical models is gaining momentum in computational epidemiology. Still, to the best of our knowledge, Sybil outperforms the available solutions in several aspects, such as its explainable and reproducible results. For instance, in^19^, the authors use a compartmental model combined with a predictive model of the pandemic to forecast its evolution 60 days in the future, building their approach on the observed data in Kenya. In particular, they propose to estimate the parameters of the compartmental model—precisely the effective reproductive number—using ARIMA and then to use the predicted values to forecast the pandemic with a SEIR compartmental model. Specifically, the authors design a compartmental model accounting for symptomatic and asymptomatic individuals, as well as mild and severe cases. Despite the many similarities, Sybil employs a much simpler compartmental model, which enables us to consider variants and still have a model without significant complexities. Also, using ARIMA makes the approach in^19^ less suitable for continuous monitoring platforms. It also makes it harder to apply the same methodology to other scenarios. Eventually, results presented in^19^ are not validated against historical data, and the forecasting accuracy is not easy to assess.

## Methods

We propose an integrated framework aimed at providing accurate and explainable predictions for epidemic spreads. Sybil combines a simple compartmental model with a machine learning-based predictive model to forecast the future progression of infection, even in the presence of multiple virus strains. At the core of Sybil, there is a simple analytical model which has a dual functionality. In the first stage of Sybil’s operation, the analytical model is used to derive the value of critical model parameters from the surveillance data. Then, these parameters’ values—ascribable to the reproductive number over time, *R*_*t*_—are used as training data for the ML component of Sybil. Based on that knowledge, it predicts the future values for the key parameters, which are then plugged back into the analytical model. Then, Sybil computes the future evolution of daily infections using the analytical model with these key parameters’ future values.

The performance evaluation of Sybil’s forecast is made by *i)* comparing it against the real data coming from the active surveillance of the pandemic situation in several European states and *ii)* comparing it against forecasts obtained by a ML approach. In this latter case, we use Prophet^21^ to predict the evolution of the different active variants in the considered period, representing a typical forecasting approach based on ML.

### Compartmental analytical model

The first component of Sybil is the analytical model. In the field of computational epidemiology, compartmental models are widely recognized and employed to study the spreading of an infection in a population. Also, compartmental models are easy to explain: the model contains several compartments, each representing a specific subpopulation, and uses rates to move individuals from one compartment to another. The simplest compartmental model to mimic the evolution of an infection in a population requires only two compartments and one rate; the two compartments represent the only two possible states of the individuals in the population—either susceptible (S) or infected (I)—and one transition rate that represents the pace at which one infected individual infects a susceptible one. Because of the two states characterizing this model, it is commonly known as the Susceptible-Infected (SI) model.

In order to capture some critical features of the SARS-CoV-2 infection, an SI model is not suitable, and a more complex one is required. Specifically, we need to model the end of the infected period explicitly, i.e., when an infected person recovers from the illness. Additionally, fatalities and reinfections must be taken into account. Hence, the compartmental model incorporated in Sybil has two additional compartments, one accounting for individuals recovered from the infection (R) and one for the deceased (D), with the corresponding rates. Specifically, the model used is a Susceptible-Infected-Recovered-Deceased-Susceptible (SIRDS) compartmental model with reinfections—represented in Figure 1-*(a)*— that is described by the system of difference equations in Equation 1. In this system, *β* represents the infection rate, *γ* the recovery rate, *λ* the fatality rate, *ν* the end-of-immunization rate, and N the total population.

**Figure 1.**
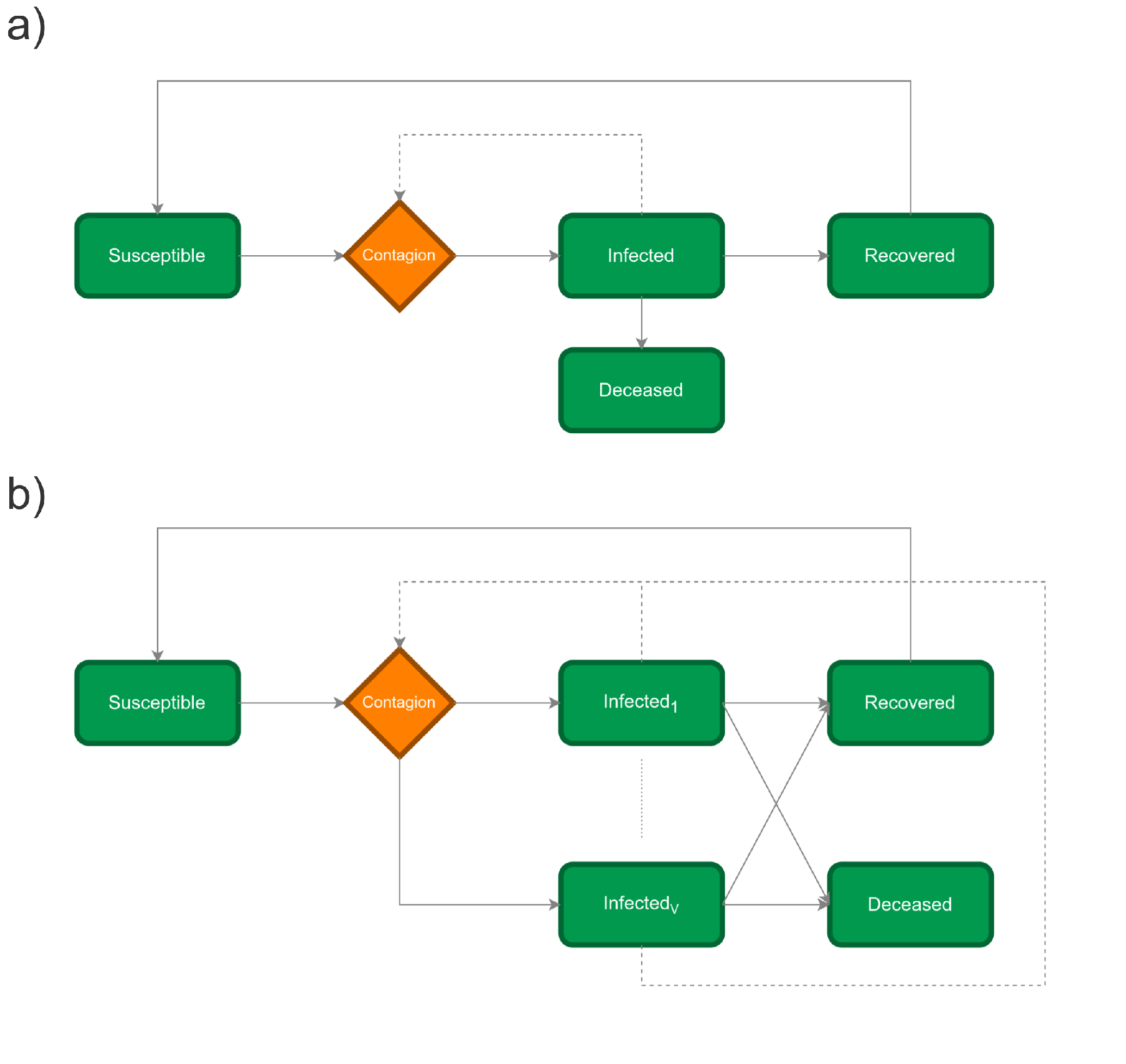
Compartmental models: *(a)* SIRDS and *(b)* SI^V^RDS. Dashed lines refer to the fact that an infection is due to contacts among susceptible and infected individuals.

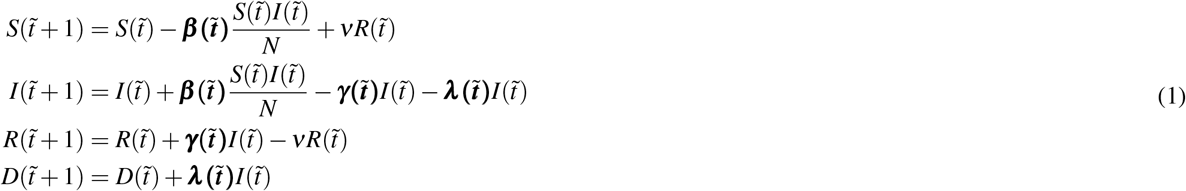

In this model, the rates are time-dependent—meaning that they may vary at each time step, with the time step corresponding to one day. The only exception is end-of-immunization rate *ν*, which is assumed to be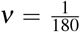, since on average the immunization due to infection is estimated to be lost after 180 days^22^.

Using the surveillance data—possibly after a pre-processing phase—, Sybil computes the evolution of the infection process by solving the SIRDS model in Equation 2, where 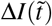 represents the new infected at time 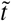, 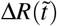 the new recoveries at time 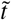 and 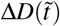 the new deceases at time 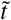 (with S(0) = N and I(0) = R(0) = D(0) = 0).

However, obtaining all the required parameters to solve equations in Equation 1 is not straightforward. Indeed, surveillance data does not provide the transition rates—namely, the bold elements in the system of difference equations—and hence a further step is required: using Equation 3 (obtained from Equation 1), we can estimate the daily infection, recovery, and fatality rates.

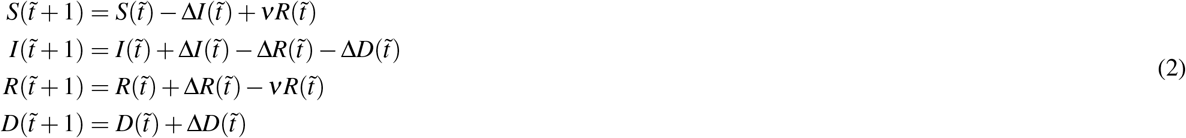

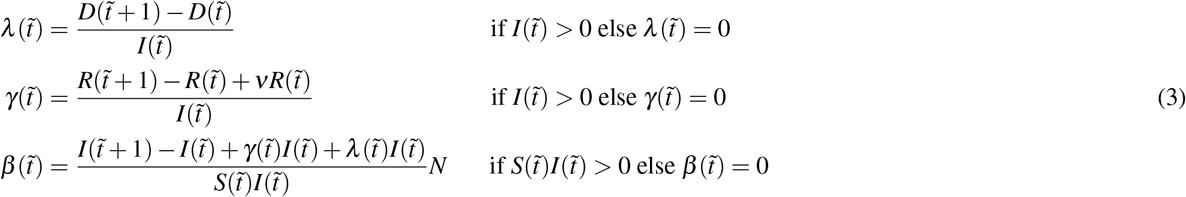

Bringing in variants into the model of Equation 1 requires introducing one additional compartment to account for infections caused by each virus strain, hence obtaining an SI^V^RDS model, where the superscript at the I stands for the maximum number of virus strains included in the model (see Figure 1-*(b)*). After introducing further compartments, also additional rates are necessary. Specifically, instead of a global infection rate 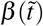, there are *V* different infection rates—one for each variant—for each time step 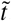 . Making a simplifying assumption, Sybil assumes that the evolution of the 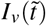 compartment—for each variant and for each time step 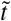 —is computed starting from the evolution of the 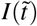 compartment of the model without variants and the daily proportion of the considered variant; that is, 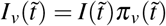, where 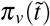 is the proportion of infections due to variant *v* at time 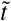 . Moreover, we assumed no correlations among variants and that being recovered from a variant *v* makes a person immune from all variants (and after 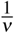 days, in average, a person will be susceptible again).

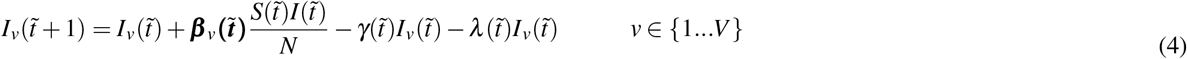

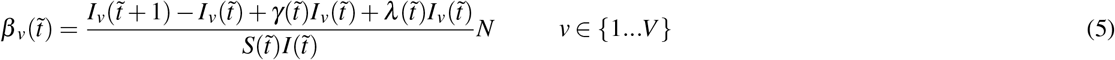

Equation 4 describes the evolution of the *I*_*v*_ compartments. Here, we used the previously computed global recovery and fatality rates, and 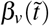 is a column vector with the time-dependent infection rates of V variants—in the supplementary material we have detailed these assumptions. Eventually, Equation 5 (obtained from Equation 4) describes how to compute the infection rates 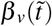 for the *v* variant.

It is worth noticing that in devising infection and fatality rates from surveillance data, we incorporate all the effects due to the different virus strains and the impact of contention policies, even if such details are not explicitly detailed in the model.

### Prophet predictive model

The second component of the Sybil framework is Prophet^21^, an open-source framework developed by Facebook for time series forecasting. It is based on an additive model design and it has been conceived to have intuitive parameters that can be adjusted without knowing the details of the underlying model. The modeling approach of Prophet combines the strengths of both statistical modeling and machine learning techniques; it utilizes a generalized additive model that incorporates piece-wise linear trends, nonlinear growth, and seasonality adjustments using a Fourier series. This flexible modeling approach enables Prophet to capture simple and complex data patterns. In particular, it consists of three main model components: trend, seasonality, and holidays combined in the following equation:

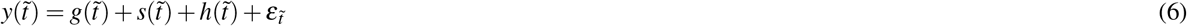

where 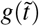 is the trend function that models non-periodic changes, 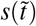 represents periodic changes (e.g., daily, weekly, and yearly seasonality), and 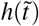 represents the effects of holidays which occur on potentially irregular schedules over one or more days. The error term 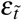 represents any characteristic changes that are not accommodated by the model. Furthermore, Prophet makes it possible to estimate uncertainty in trend forecasts. It employs Markov Chain Monte Carlo (MCMC) to generate many plausible future trajectories. The MCMC procedure randomly samples from the posterior distribution of the model parameters, allowing for a range of possible outcomes. These sampled parameter sets are then used to generate multiple forecast trajectories.

### Data

All datasets used in this work are open source and publicly available. The surveillance data used for Italy and Austria in the *Results* section are available in the COVID19 R library^23,24^ (for the results presented in the *Results* section we used a snapshot dated November 22^nd^, 2023). This data comprises all the data reported in Figures 2 to 6 plus other data not relevant to the present study—such as the number of vaccinations, tests, hospitalizations and people in intensive care, information about applied countermeasures, data on mobility, etc. In particular, from the COVID-19 Data Hub^23,24^, we used the following data (from February 2020 to May 2023): the cumulative number of cases, the cumulative number of recoveries, the cumulative number of deaths, and the total population. Starting from these, we computed the daily new cases, the daily new recoveries, and the daily new deaths we used in Equation 2.

**Figure 2.**
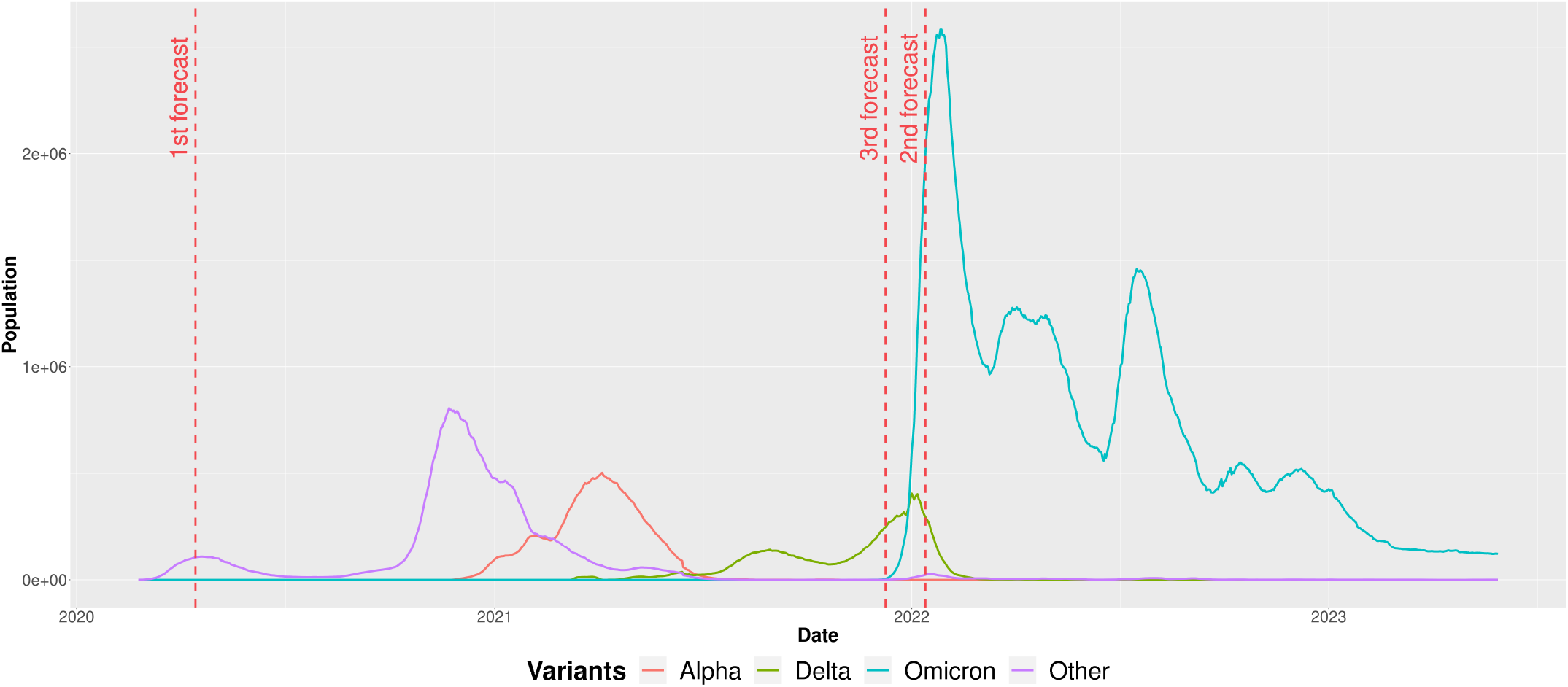
Daily active cases in Italy from February 2020 to May 2023 for the four main SARS-CoV-2 strains. Vertical dashed lines mark the selected dates to test Sybil’s forecasting.

Information about active virus strains comes from the European Center for Disease Control (ECDC), specifically, the one reported in^25,26^ (for the results presented in the *Results* section we used a snapshot dated July 25^th^, 2023). Here, ECDC collected the result of serological tests, and there is information about a significant number of variant lineages, such as B.1.1.7, BA.1, BA.2, P.1, XBB, and many others. To use such data, we aggregated these lineages in four main variant families with the aim of using the WHO variants labels: Alpha, Delta, Omicron, and *Other*^27^. In particular, the *Other* variant comprises the initial SARS-CoV-2 lineage, all the other lineages (e.g., Beta, Gamma, Kappa), and some noisy values in the surveillance data. Since variants’ diffusion data is provided weekly, in a pre-processing phase, we expanded such data to devise approximated daily values. Specifically, we employed splines—piece-wise-defined mathematical functions that use multiple polynomial segments to create a smooth and continuous curve—to obtain daily values.

In the following section, we used time-dependent recovery rates thanks to data availability for the selected countries— namely Italy and Austria. However, surveillance data available for many countries worldwide does not report data to devise daily recovery rates. Hence, to dispense with data on recoveries, in the supplementary material we report results obtained with fixed recovery rate with mean 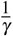 equals to 14 days—same for each variant. The results obtained are promising and show the robustness of the Sybil approach.

## Results

This section starts focusing on surveillance data from Italy and spanning a period from February 2020 to May 2023. During this period, there are a significant number of lineages, some coexisting and others with an evolutionary advantage taking over. In Figure 2, lineages are aggregated into the four main variants introduced above.

Accurate forecasts strongly depend on the regularity of the data to predict: it is pretty easy to foresee the future evolution of some dimensions increasing—or equally decreasing—linearly. Conversely, predicting the behavior in the presence of sudden changes in those quantities is much harder. Unfortunately, outbreaks, as well as peak infection fading, exhibit such a behavior. Vertical dashed lines in Figure 2 mark the time point picked to evaluate the accuracy of Sybil. The first two selected points are placed in the ascending part of an outbreak but close enough to the peak for accurate predictions, Sybil must successfully reproduce a change in the concavity of the function. The third selected point is placed just before the start of Omicron’s outbreak to show that Sybil is also able to predict a new emerging variant/outbreak. Further experiments and scenarios are available in the supplementary material.

The first forecast scenario corresponds to the first infection wave, which in Italy started in February 2020. Only one SARS-CoV-2 strain was detected during this initial wave, namely the *Other* variant of Figure 2. Figure 3-*(a)* reports four different forecasts, all starting on April 15^th^ 2020 and spanning several forecast windows—from one to four weeks. For each forecasting window, we compare predictions obtained using Sybil (green line) and the standard approach used in the literature (red line), which requires using the selected forecasting approach—Prophet in our case—to project the number of infections in the coming weeks. The two approaches use the same period as training data (black line) for a fair comparison. They are compared and contrasted against the surveillance data for the period spanning the forecasting window (blue line). Comparing predictions obtained with the plain use of Prophet to forecast the evolution of the number of daily infections, we notice that it fails to predict the plateau characterizing the highest part of the peak. Considering Sybil’s predictions, we see that starting from short-term predictions—i.e., seven days—it catches the decreasing trend. Increasing the forecasting window, Sybil always provides an excellent approximation of the future evolution of the daily infections, but for four weeks. In this latter case, Sybil’s predictions diverge in magnitude from the ground truth, but not in trend. It provides a qualitative indication of the future evolution of the pandemic, which eventually fades. Figure 3-*(b)* shows the infection rate (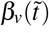 for the observation period. Here, we clearly see that the values for the infection rate are noisy; what we see is not a smooth line, but saw-tooth function with many peaks and valleys. Nonetheless, the range of variation of that function is limited and with a clear trend, which Sybil easily learns and replicates.

**Figure 3.**
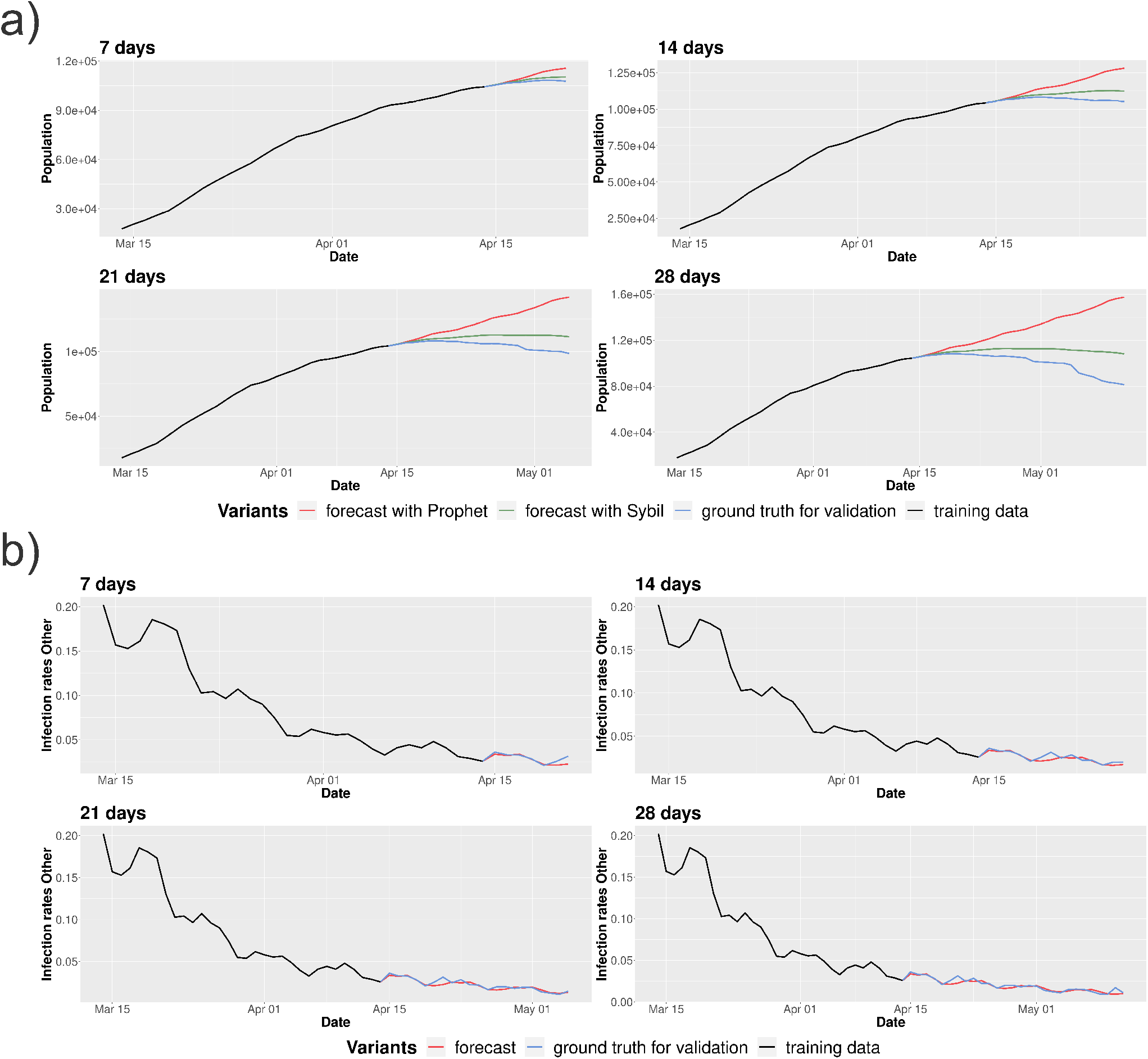
The figures refer to the first scenario in which we forecast starting from April 14^th^ 2020. Figure *(a)* shows the evolution of infections using Sybil (green line) and Prophet (red line) using the same period as training data (black line) comparing and contrasting the predictions against the surveillance data for the period spanning the forecasting window (blue line). Figure *(b)* shows Sybil applied on the infection rate *β*_*v*_(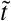) of the *Other* variant (the red line shows the prediction, while black and blue lines represent the training data and the ground-truth values extracted from the surveillance data, respectively).

The second scenario considers a period spanning the highest peak in the data set. Here, we consider the period from December 13^th^ 2021 to January 13^th^ 2022 as training data, and we forecast the daily infections for the period January-February 2022 (starting from January 14^th^). In Italy, there were three active variants within this time window: Omicron, Delta, and the *Other* variant. Figure 4-(a) shows the number of daily infections for the three active variants and compares the ground truth (dashed line) with Sybil’s forecasts for one to four weeks. Again, forecasts are highly accurate for seven to twenty-one days long predictions and slightly anticipate the peak’s descendent phase at four weeks. In Figure 4-*(b)*, the two approaches are contrasted against the ground truth. Like the first scenario, the Prophet’s predictions do not capture the peak. Not only do Prophet’s predictions get far from the real data—they grow while the infection fades—but, in this scenario, they do not even provide a valid qualitative prediction.

**Figure 4.**
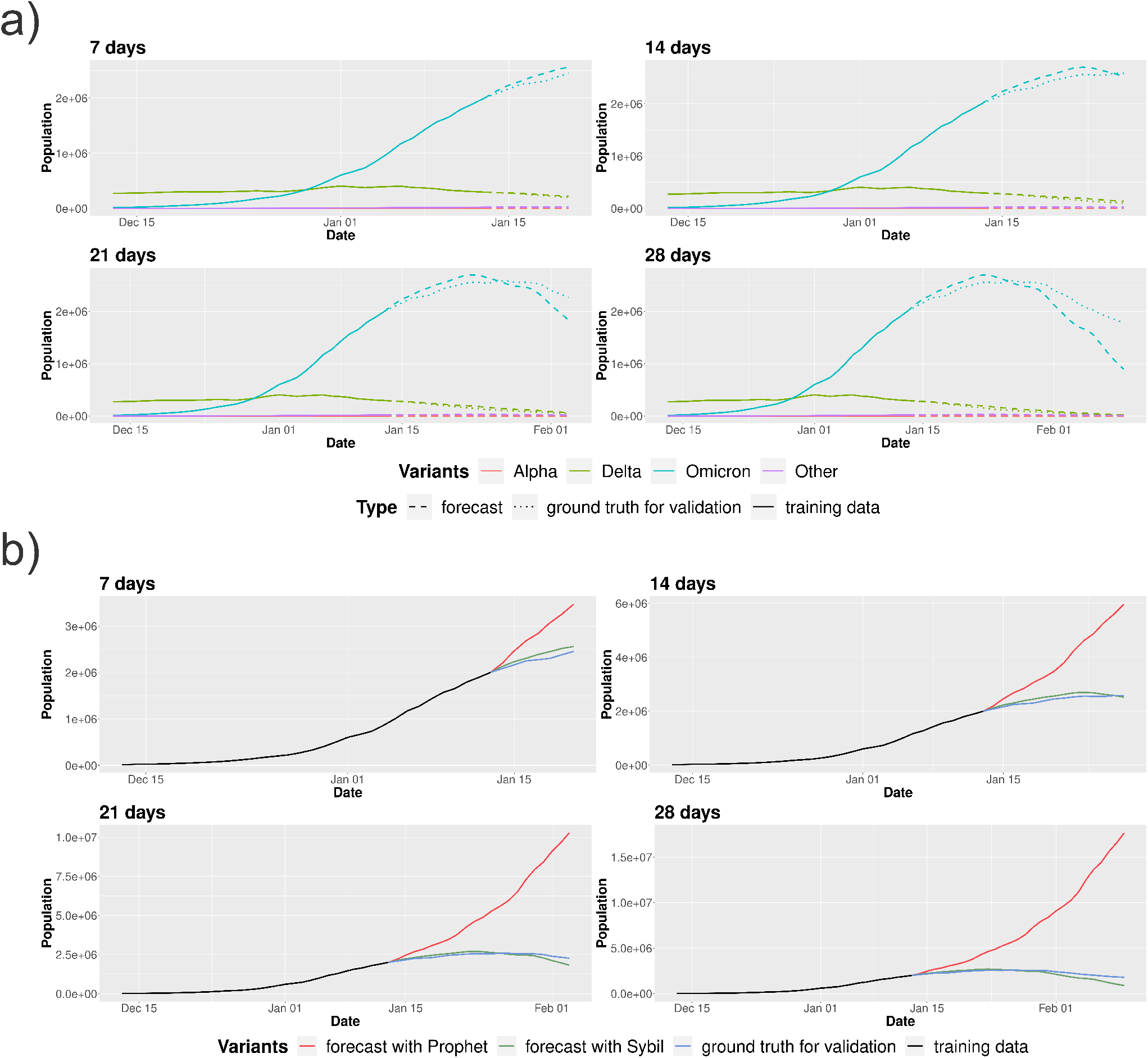
The figures refer to the second scenario in which we forecast starting from January 13^th^ 2022. Figure *(a)* show the evolution of infections using Sybil (the dashed line shows the prediction, while solid and dotted lines represent the training data and the ground-truth values extracted from the surveillance data, respectively). Figure *(b)* shows the comparison between Sybil (green line) and Prophet (red line) on the number of infections for the Omicron variant using the same period as training data (black line) comparing and contrasting the predictions against the surveillance data for the period spanning the forecasting window (blue line).

The third point picked to test Sybil’s predictions falls just before the explosion of the largest outbreak caused by the Omicron variant. In particular, Figure 5 shows how the one-week forecast changes moving the training window from December 18^th^, 2021 to December 27^th^, 2021. In this scenario, the ascending trajectory is very steep, and Sybil finds it harder to calibrate with respect to the previous cases. Here, the Omicron variant is a new emerging variant and Sybil initially foresees a more aggressive exponential growth. Looking at Figure 5 we can state that Sybil nailed the qualitative prediction, but needed more data to calibrate it. We can also see that Sybil captures well the prediction on the other active variant—the Delta variant. Here we are showing the best results obtained in this scenario. In the supplementary material we provide more details on this scenario and show another case where an explosive outbreak occurs, discussing the convenience of establishing a continuous monitoring system.

**Figure 5.**
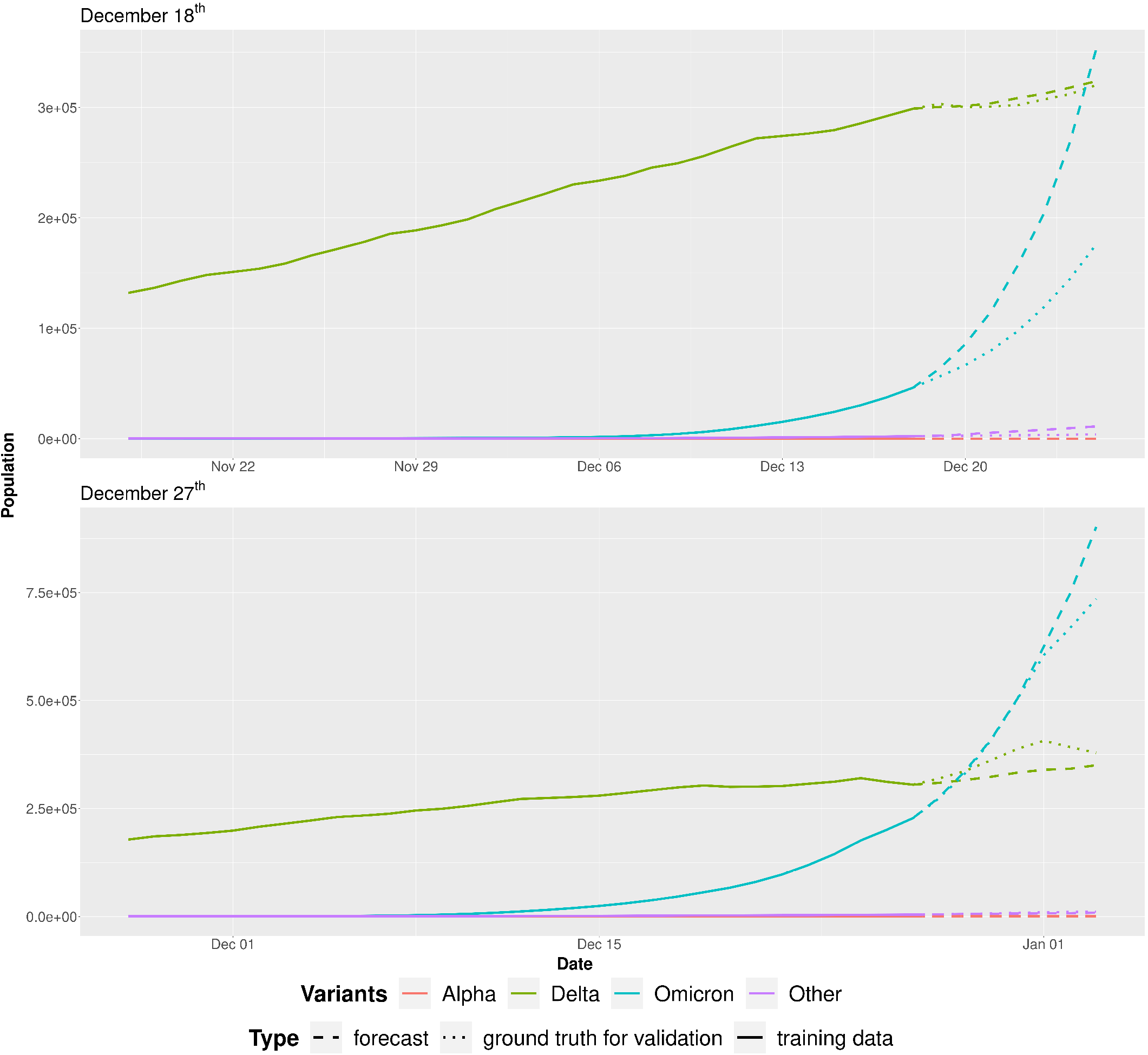
Evolution of infections using Sybil on the third scenario. In the first plot we forecast starting from December 18^th^, 2021, while in second one we forecast starting from December 27^th^, 2021. Both the plots refer to a forecast one week into the future. The dashed line shows the prediction, while solid and dotted lines represent the training data and the ground-truth values extracted from the surveillance data, respectively.

Sybil provides a robust, easily portable approach to different use cases without requiring modifications. For instance, in Figure 6 we apply Sybil to surveillance data from Austria. Figure 6-*(a)* shows the daily infections for the different SARS-CoV-2 strains active in the period from February 2020 to May 2023. As in the case of Italy, lineages are grouped into four main virus strains—Alpha, Delta, Omicron, and the *Other* variant. In Figure 6-*(b)*, data from January 1^st^ 2022 to February 1^st^ 2022 are employed to train the models, and we compare forecasts obtained for February 2022 (starting from February 2^nd^). During this period, there were two active variants: Omicron and the *Other* variant, as in the second scenario considered for Italy. The initial point chosen for the forecasts is again placed in the surroundings of the highest peak so that correctly predicting the future evolution of the infection is much more challenging. Nonetheless, Sybil shows high accuracy in predicting the future trajectory of the infection; despite a slightly different evolution, Sybil correctly predicts the diffusion slowdown, the successive decreasing phase. Eventually, prediction and surveillance data meet at the end of the four-week-long forecast.

**Figure 6.**
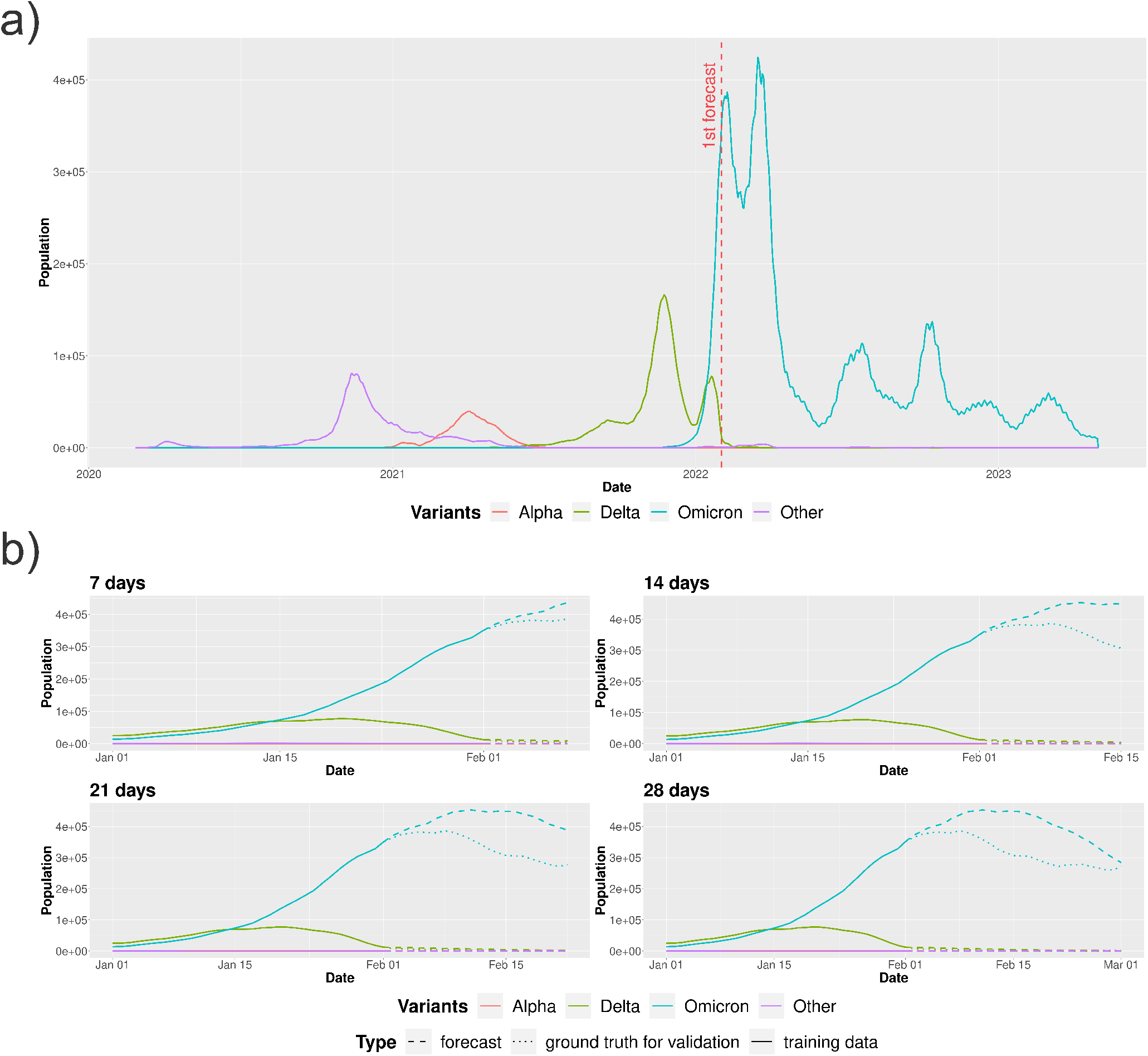
The figures show *(a)* the evolution of infections from February 2020 to May 2023 in Austria with the considered forecast scenario and *(b)* the evolution of infections using Sybil on the considered scenario in which we forecast starting from February 1^st^ 2022 (the dashed line shows the prediction, while solid and dotted lines represent the training data and the ground-truth values extracted from the surveillance data, respectively).

## Discussion

ML approaches have already shown their power when dealing with complexity, thanks to their excellent capabilities in exploiting non-trivial correlations often inaccessible with other tools. Despite the many advantages of employing ML approaches, there are also some caveats worth noting. First, most of these approaches require a tremendous amount of data to learn from, and data available from the surveillance might not be enough for most ML approaches. Sybil faces this issue from two different standpoints; it employs Prophet, a hybrid approach, using ML techniques in combination with simulations—refer to the *Prophet predictive model* subsection for further details. On the other hand, Sybil does not face the challenge of forecasting the virus spread as a single task. First, it tries to predict the rates and then computes the future dynamics using a compartmental model. Therefore, Sybil, combining Prophet and compartmental models, exploits the two approaches in the respective field of application—where they perform the best—and strengthens the individual weaknesses. Specifically, by providing the compartmental model with parameters extracted from the real data or forecasts, there is no need to tune the model and estimate the missing parameters. Estimating model’s parameters is a resource-demanding and time-consuming task. Also, it is an activity tightly related to the specific situation or scenario under evaluation. This makes the model almost impossible to apply in a different setting without estimating the parameters again. Hence, Prophet makes Sybil’s approach easily deployable in new scenarios without requiring additional tasks, but the data must comply with the daily requirement. Likewise, the use of compartmental models in predicting the future trend of the infection makes results more straightforward to explain, for instance, to policymakers, who often do not trust predictions without a robust interpretation.

In the *Results* section, we presented Sybil’s forecasts of different lengths for several European states and periods spanning from February 2020 to May 2023. To showcase Sybil’s capabilities, all the forecasting periods cover important changes in the first derivative of the number of daily infections. Then, Sybil’s predictions are contrasted against the surveillance data and the plain application of Prophet. Results shows the superiority of Sybil’s approach on the plain forecast of the number of daily infections; in particular, Sybil outperforms the plain application of Prophet when primary changes in the virus diffusion happens. For instance, Figure 4-*(b)* clearly shows the added value of Sybil approach: it predicts with great accuracy the peak of the current wave while Prophet alone fails to predict the decreasing phase of the infection peak. Further experiments and scenarios are available in the supplementary material. Here, for instance, some experiments made choosing forecasting periods far from peaks show that similar and accurate forecasting can be obtained using both approaches and new variants rising or new outbreaks can be predicted accurately using Sybil to set up a continuous monitoring system.

The presented methodology is very close to being a continuous monitoring system, but strongly depends on the availability of data. In particular, in^23,24^ there are few nations with a complete time series (with a daily step) for the data we used (i.e., cumulative number of cases, cumulative number of deceases, cumulative number of recoveries). Possible extensions to this methodology would be to use a fixed recovery rate—same or different for each variant—to dispense with data on recoveries which are often not available—in the supplementary material we show some results related to this extension—and to include a pre-processing step using a technique (e.g., splines) to fill missing data and to move from a weekly to a daily step in case of availability of data with a weekly step—without alter too much the information present in the time series.

## Conclusion

The global COVID-19 pandemic has brought to light the urgent necessity for sophisticated tools capable of monitoring and predicting the trajectory of infections within the population. This paper introduces a cutting-edge framework designed for continuous monitoring and forecasting, seamlessly integrating machine learning-based predictive models with compartmental models.

Sybil distinguishes itself by delivering forecasts that are not only reliable but also readily explainable, as evidenced by thorough experimental validation. The adaptability of this innovative approach is clearly demonstrated through its successful application to diverse surveillance data from two European countries, specifically Italy and Austria.

By integrating data-centric and analytic approaches, Sybil effectively addresses the inherent limitations of each method. This amalgamation not only enhances the tool’s ability to forecast the evolution of COVID-19 but also positions Sybil as a versatile instrument for predicting the trajectory of various other diseases, thus broadening its scope and impact in the field of infectious disease modeling.

## Data Availability

COVID-19 data used in this study are available in the COVID19 R library and in the ECDC variants data.
The framework developed for the analysis presented in this work is available at https://github.com/daniele-baccega/sybil-forecasting. To reproduce the results, see supplementary material.

https://cran.r-project.org/web/packages/COVID19/index.html

https://www.ecdc.europa.eu/en/publications-data/data-virus-variants-covid-19-eueea

https://github.com/daniele-baccega/sybil-forecasting

## Acknowledgements

D.B. is a Ph.D. student enrolled in the National Ph.D. in Artificial Intelligence, XXXVII cycle, health and life sciences course organized by Università Campus Bio-Medico di Roma.

## Author contributions statement

D.B. discussed, developed and tested the framework, conceived and conducted the experiments. P.C., A.F.A. and M.S. discussed and designed the framework, conceived the experiments. All authors reviewed the manuscript.

## Additional information

### Availability of materials and data

COVID-19 data used in this study are available in the COVID19 R library^23,24^ and in the ECDC variants data^25,26^. The framework developed for the analysis presented in this work is available at https://github.com/daniele-baccega/sybil-forecasting. To reproduce the results, see supplementary material.

