## Supplementary Material for "Enhancing COVID-19 Forecasting Precision through the Integration of Compartmental Models, Machine Learning and Variants"

#### Contents

**1 Introduction** 1

**2 SIRDS compartmental model** 1

**3 Why Prophet?** 2

**4 Other results on Italy** 2

4.1 Results with V $=4$ . . . 2

Relative errors • Fixed recovery rate

4.2 Results with V $=11$ . . . 17

**Other results on Austria** 24

**5 Continuous monitoring** 31

**6 How to reproduce the results** 35

6.1 Requirements . . . 35

6.2 Reproduce . . . 36

#### 1 Introduction

In this document we show more details on the SIRDS compartmental model, the reason behind Prophet's choice and other results related to Italy and Austria with the same aggregation of lineages introduced in the main document ($V = 4$). Moreover, we show some results related to Italy with a different aggregation of lineages ($V = 11$) and we discuss about the implementation of a continuous monitoring system. Finally, we show how to reproduce all the results presented in the paper.

#### 2 SIRDS compartmental model

In the main document we directly presented the discretization of the used compartmental model. However, it is essential to note that, by definition, a Susceptible-Infected-Recovered-Deceased-Susceptible (SIRDS) compartmental model is described by the system of Ordinary Differential Equations (ODEs) in Equation SM1.

$$
\begin{aligned}
\frac{dS}{dt} &= -\beta(t)\frac{S(t)I(t)}{N} + \nu R(t) \\
\frac{dI}{dt} &= \beta(t)\frac{S(t)I(t)}{N} - \gamma(t)I(t) - \lambda(t)I(t) \\
\frac{dR}{dt} &= \gamma(t)I(t) - \nu R(t) \\
\frac{dD}{dt} &= \lambda(t)I(t)
\end{aligned}
\tag{SM1}
$$

We discretized this continuous system of ODEs obtaining a system of difference equations that express how the sizes of the compartments vary over time on a discrete scale – Equation SM2; $t$ is a continuous variable, while $\tilde{t}$ is a discrete one that takes as values multiples of $\Delta\tilde{t}$.

$$
\begin{aligned}
\frac{S(\tilde{t}+\Delta\tilde{t})}{\Delta\tilde{t}} &= S(\tilde{t}) - \beta(\tilde{t})\frac{S(\tilde{t})I(\tilde{t})}{N} + \nu R(\tilde{t})\\
\frac{I(\tilde{t}+\Delta\tilde{t})}{\Delta\tilde{t}} &= I(\tilde{t}) + \beta(\tilde{t})\frac{S(\tilde{t})I(\tilde{t})}{N} - \gamma(\tilde{t})I(\tilde{t}) - \lambda(\tilde{t})I(\tilde{t})\\
\frac{R(\tilde{t}+\Delta\tilde{t})}{\Delta\tilde{t}} &= R(\tilde{t}) + \gamma(\tilde{t})I(\tilde{t}) - \nu R(\tilde{t})\\
\frac{D(\tilde{t}+\Delta\tilde{t})}{\Delta\tilde{t}} &= D(\tilde{t}) + \lambda(\tilde{t})I(\tilde{t})
\end{aligned}
\tag{SM2}
$$

The rates (except for the end-of-immunization rate) are time-dependent and vary at each time step – with the time step corresponding to one day, i.e. $\Delta\tilde{t} = 1$, which is the frequency of the observations in the datasets – Equation SM3.

$$
\begin{aligned}
S(\tilde{t}+1) &= S(\tilde{t}) - \beta(\tilde{t})\frac{S(\tilde{t})I(\tilde{t})}{N} + \nu R(\tilde{t})\\
I(\tilde{t}+1) &= I(\tilde{t}) + \beta(\tilde{t})\frac{S(\tilde{t})I(\tilde{t})}{N} - \gamma(\tilde{t})I(\tilde{t}) - \lambda(\tilde{t})I(\tilde{t})\\
R(\tilde{t}+1) &= R(\tilde{t}) + \gamma(\tilde{t})I(\tilde{t}) - \nu R(\tilde{t})\\
D(\tilde{t}+1) &= D(\tilde{t}) + \lambda(\tilde{t})I(\tilde{t})
\end{aligned}
\tag{SM3}
$$

In Equation (4), in the main document, we took infections from the $I(\tilde{t})$ compartment instead of from the $I_v(\tilde{t})$ compartments. This is necessary to avoid approximation errors since we could have, especially in an early stage of a variant, small values inside the $I_v(\tilde{t})$ compartments that could create some problems with Sybil. In this way we have a vector of rates $\beta_v(\tilde{t})$ and not a matrix. Moreover, even if we would use a matrix we could not compute most of the values – we can compute only the values on the main diagonal. To compute all the values we would need to distinct the recoveries and the deaths for each variant (and, therefore, the recovery and fatality rates), but we do not have these ground-truth data inside the datasets we used – and, more generally, these information are rarely available.

### 3 Why Prophet?

We chose Prophet as machine learning-based predictive model for its flexibility, non-linearity, robustness, and user-friendliness. We also conducted a comparative analysis between Prophet and ARIMA [SM1]. Specifically, we performed preliminary work using ARIMA, which resulted in comparable outcomes to those achieved with Prophet. However, it is important to note that ARIMA requires a substantial amount of domain knowledge to effectively select and interpret parameters, such as the maximum orders of differencing, auto-regressive components, moving average components, and others. Moreover, ARIMA is susceptible to significant trend errors when there are changes in trend near the cutoff period and may struggle to capture seasonality [SM2].

### 4 Other results on Italy

#### 4.1 Results with V $= 4$

Figure SM1 shows the evolution of the $I(\tilde{t})$ compartment from February 2020 to May 2023 in Italy obtained using Equation (2) (see *Compartmental analytical model* section in the main document) with the three previously considered scenarios (first, second and third) and with other three new scenarios (fourth, fifth and sixth). Figures SM2, SM3 and SM4 show the evolution of infection, recovery and fatality rates respectively, in the same period, extracted using Equation (3). Figures SM5 and SM6 show the daily proportion of each variant and the evolution of the $I_v(\tilde{t})$ compartments obtained using Equation (4), respectively. Figure SM7 shows the evolution of infection rates for each variant.

In particular, from these figures we can state that there is a clear correlation between the infection rate for each variant $v$ and the evolution of the $I_v(\tilde{t})$ compartment in Figure SM6. Moreover, we can see that the recovery rate changes with respect to variants and that the fatality rate seems to be stable – after an initial transient period. From these figures it is clear that the range of variation of the rates is much smaller than the infected population one, which is one of the reasons why Sybil works better with respect to the plain use of Prophet.

Figures SM8, SM9 and SM10 show the prediction obtained applying Sybil on the global infection, recovery and fatality rates in the first scenario.

The fourth scenario falls in the middle of the Summer 2022. In this period, Italy observed a recrudescence of the pandemic. Omicron is the primary strain responsible for this peak. In this latter case, we consider the period from June 14$^{\text{th}}$ 2022 to July

14th 2022 as training data, and we forecast on July-August 2022 (starting from July 15th). Again, there is a slope change in the Omicron trajectory – although not as intense as in the second scenario – and Sybil captures it very well, see Figure SM11. In this particular scenario, Sybil performs surprisingly well, and the forecast accuracy does not degrade when moving forward in the future.

In the fifth scenario, we used the period from May 30th 2020 to June 30th 2020 as training data, and we forecasted on July 2020 (starting from July 1st). In this scenario, only one active variant (the *Other* variant) exists. In this period, we do not have a drastic change in the slope of the trajectory, and in Figure SM12 we can see that both the approaches are able to capture this less sudden change of slope, but again Sybil is slightly better than the plain application of Prophet.

In the sixth scenario, we used the period from January 7th 2023 to February 7th 2023 as training data, and we forecasted on February-March 2023 (starting from February 15th). Here there are two active variants: Omicron and the *Other* variant. Here the trajectory is decreasing, and we capture almost perfectly the future trend – Figure SM13.

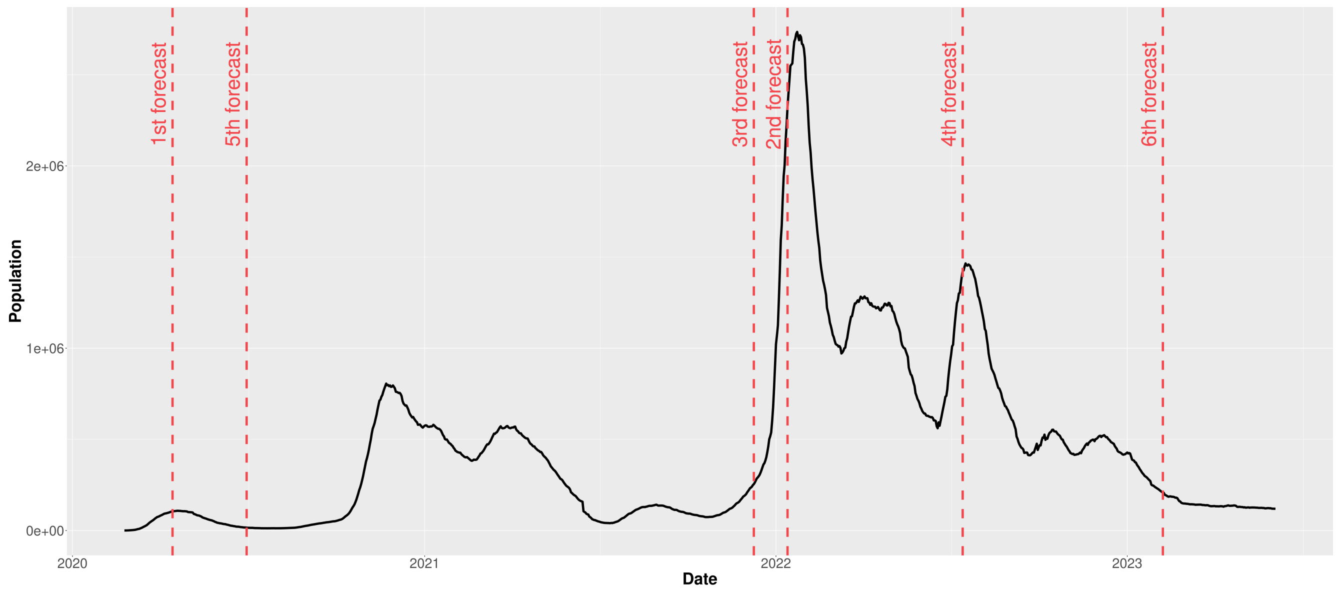


**Figure SM1.** Evolution of the $I(\tilde{t})$ compartment from February 2020 to May 2023 in Italy with the six considered forecast scenarios. Vertical dashed lines mark the selected dates to test Sybil's forecasting.

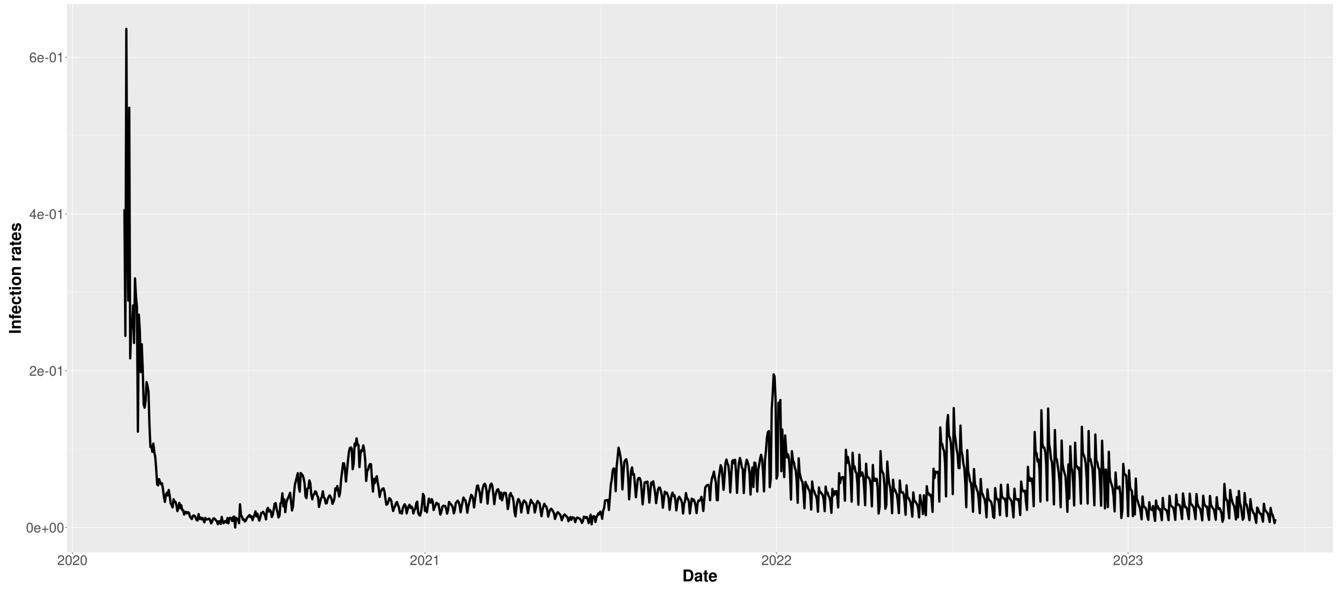


**Figure SM2.** Evolution of the infection rates $\beta(\tilde{t})$ from February 2020 to May 2023 in Italy.

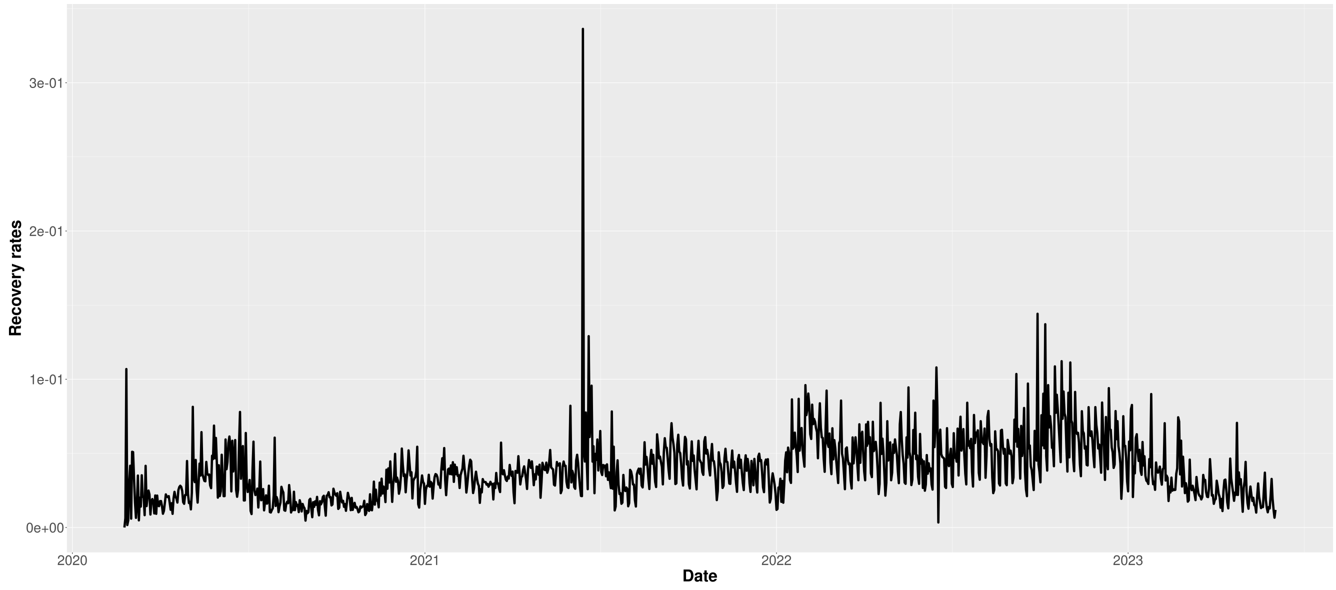


**Figure SM3.** Evolution of the recovery rates $\gamma(\tilde{t})$ from February 2020 to May 2023 in Italy.

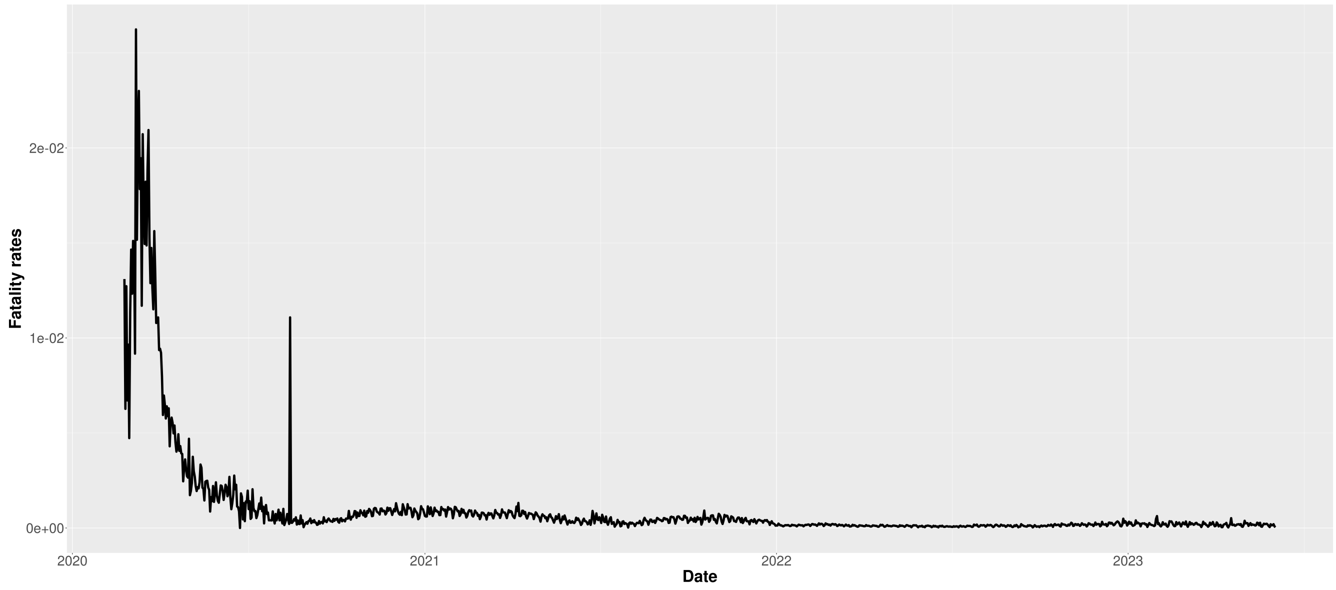


**Figure SM4.** Evolution of the fatality rates $\lambda(\tilde{t})$ from February 2020 to May 2023 in Italy.

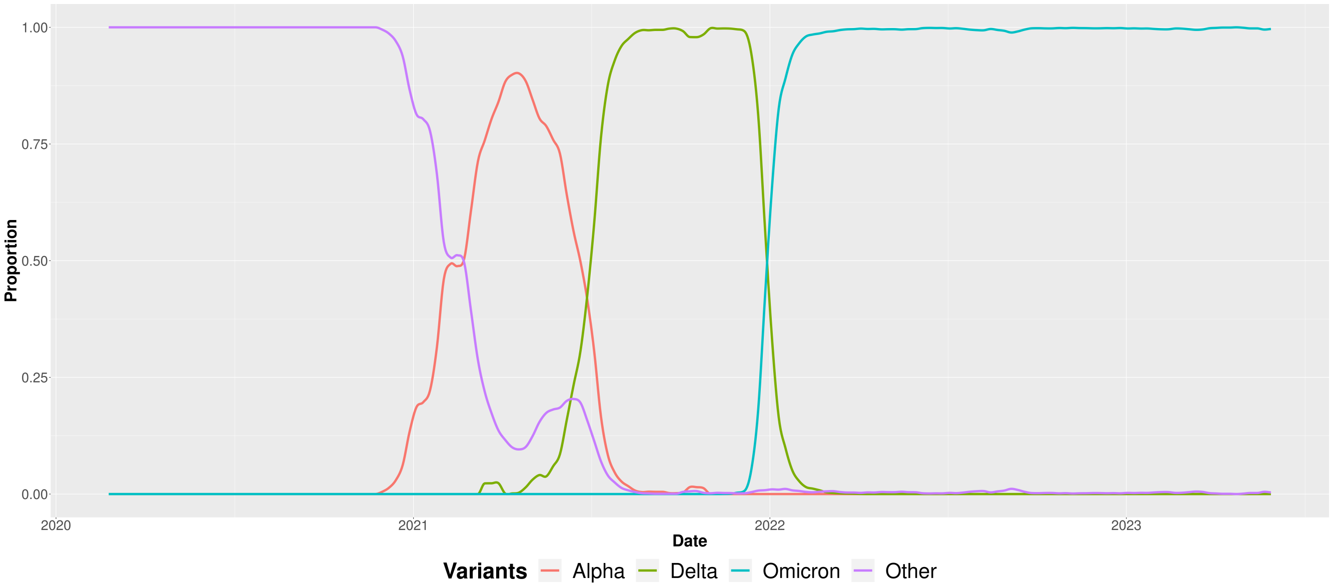


**Figure SM5.** Proportion of the aggregated variants from February 2020 to May 2023 in Italy.

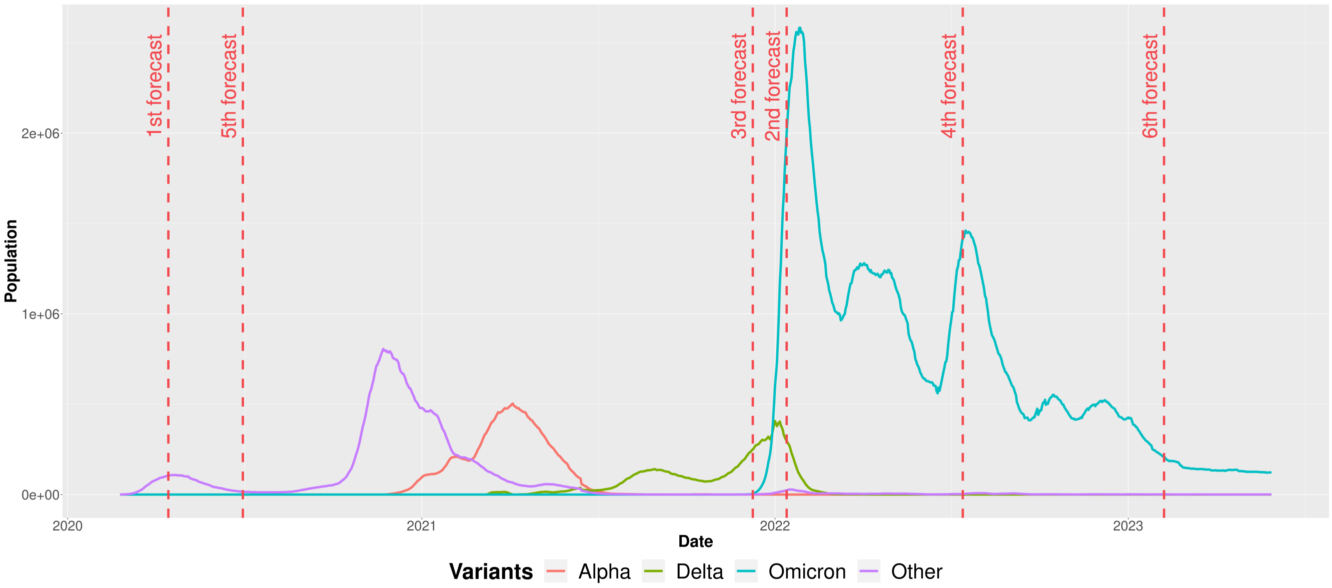


**Figure SM6.** Evolution of the $I_v(\tilde{t})$ compartments from February 2020 to May 2023 in Italy with the six considered forecast scenarios. Vertical dashed lines mark the selected dates to test Sybil's forecasting.

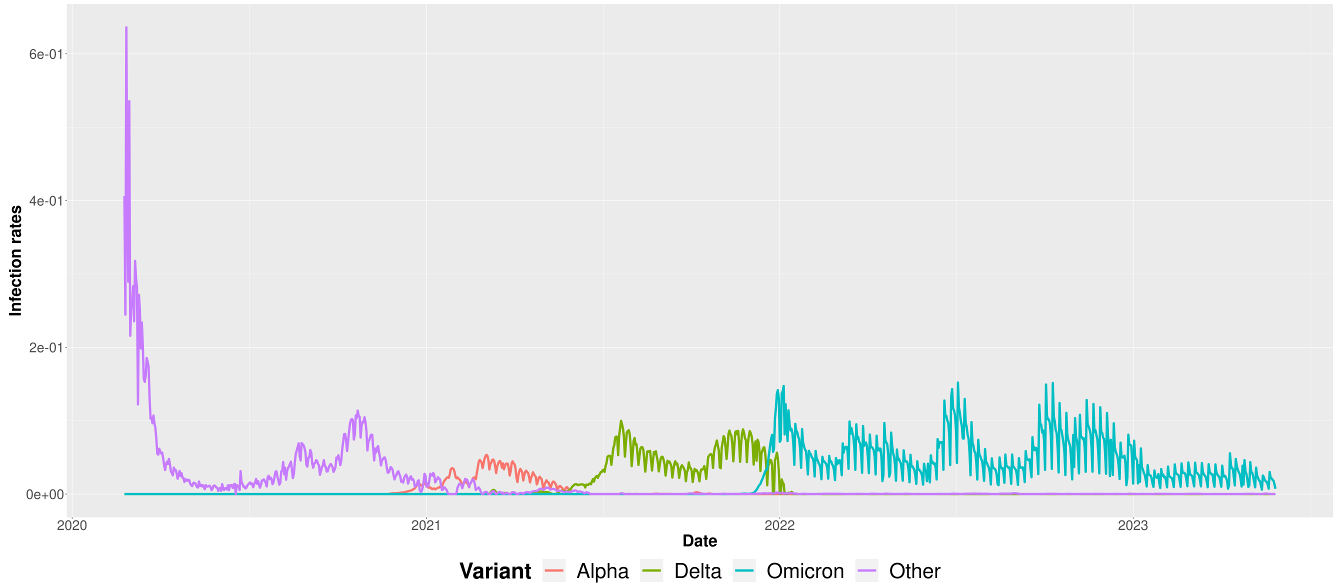


**Figure SM7.** Evolution of the infection rates $\beta_v(\tilde{t})$ for each variant from February 2020 to May 2023 in Italy.

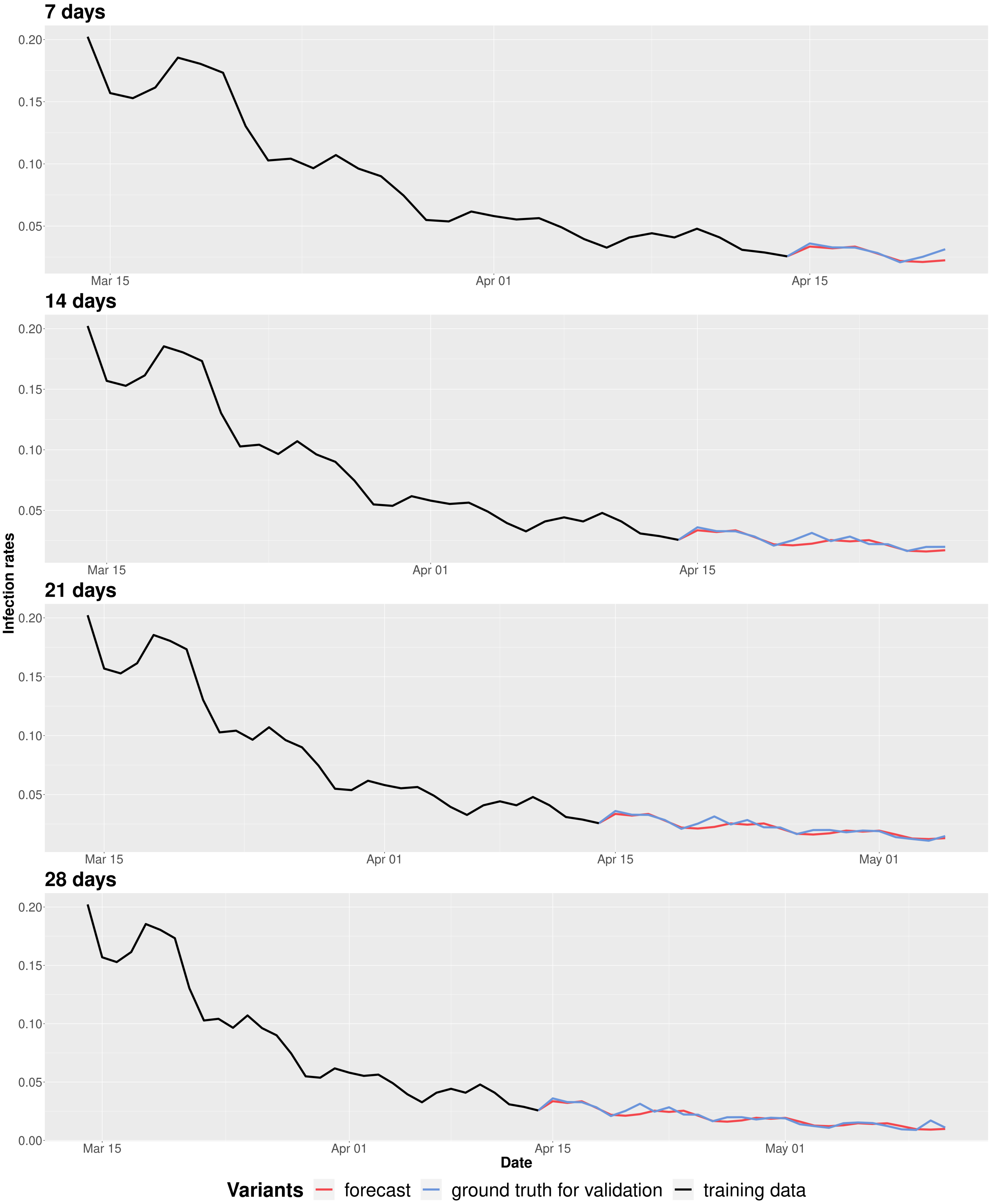


**Figure SM8.** Sybil applied on the global infection rates $\beta(\tilde{t})$ of the first scenario in which we forecast starting from April 14$^{th}$ 2020 (the red line shows the prediction, while black and blue lines represent the training data and the ground-truth values extracted from the surveillance data, respectively).

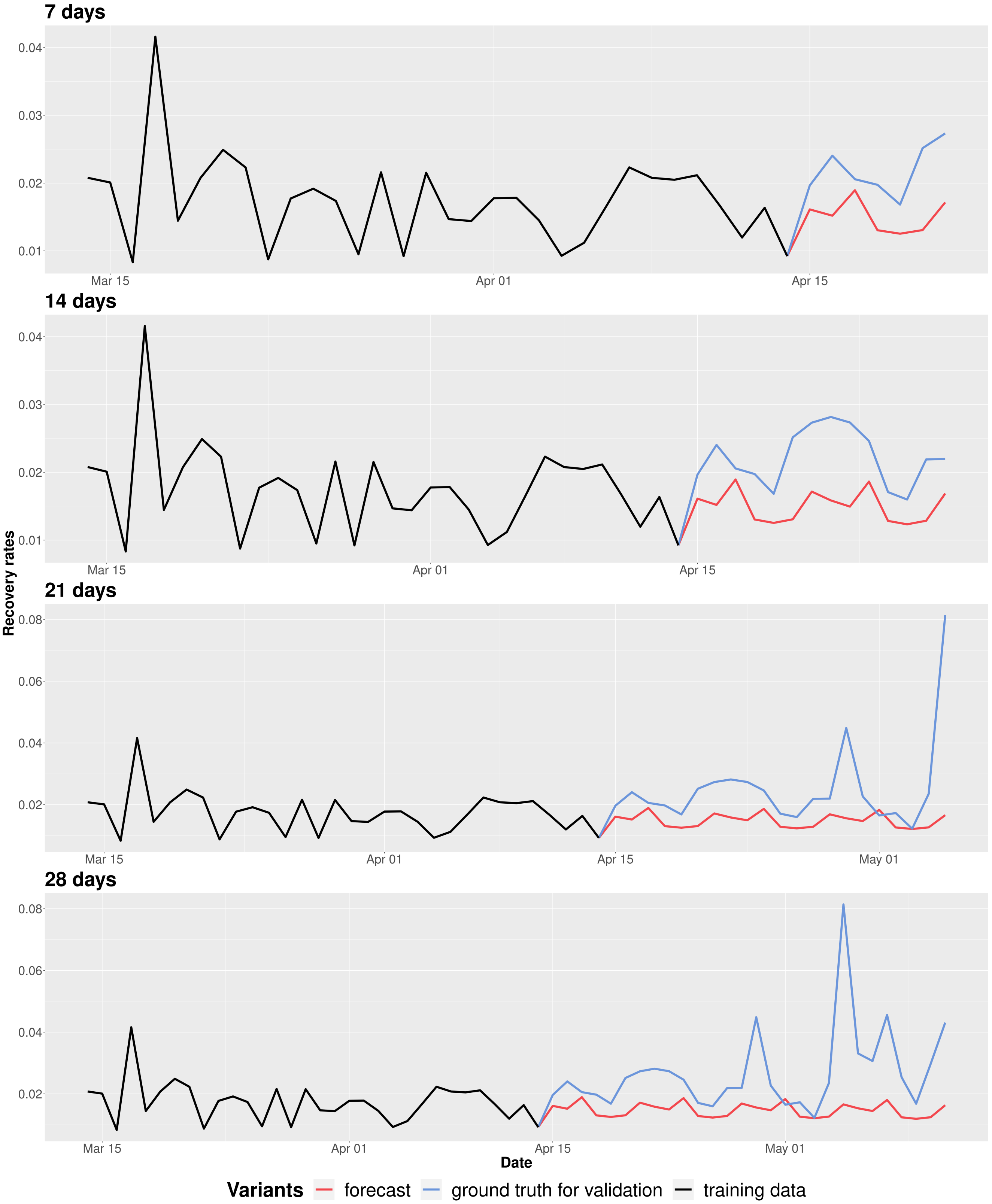


**Figure SM9.** Sybil applied on the global recovery rates $\gamma(\tilde{t})$ of the first scenario in which we forecast starting from April 14$^{th}$ 2020 (the red line shows the prediction, while black and blue lines represent the training data and the ground-truth values extracted from the surveillance data, respectively).

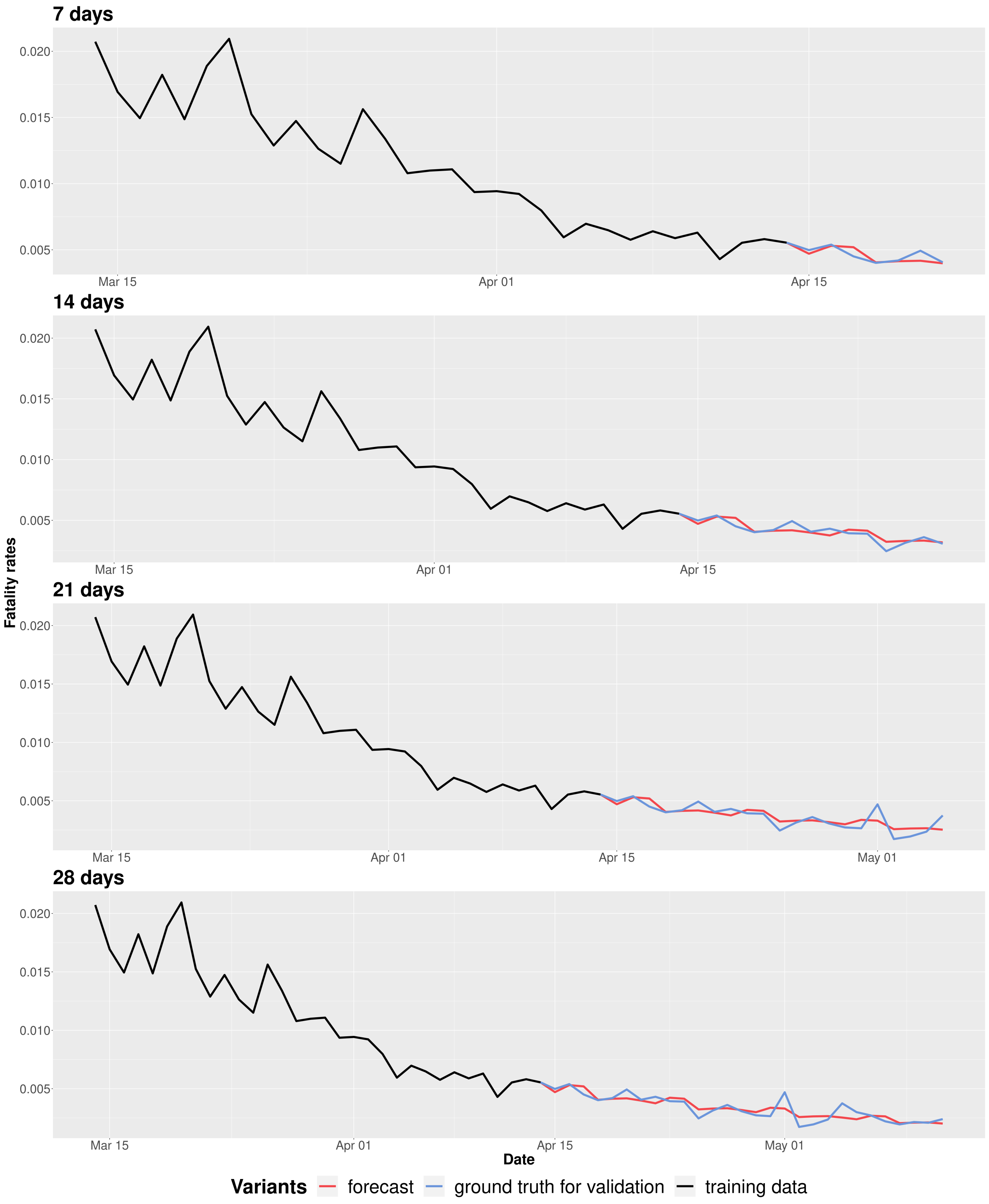


**Figure SM10.** Sybil applied on the global fatality rates $\lambda(\tilde{t})$ of the first scenario in which we forecast starting from April 14th 2020 (the red line shows the prediction, while black and blue lines represent the training data and the ground-truth values extracted from the surveillance data, respectively).

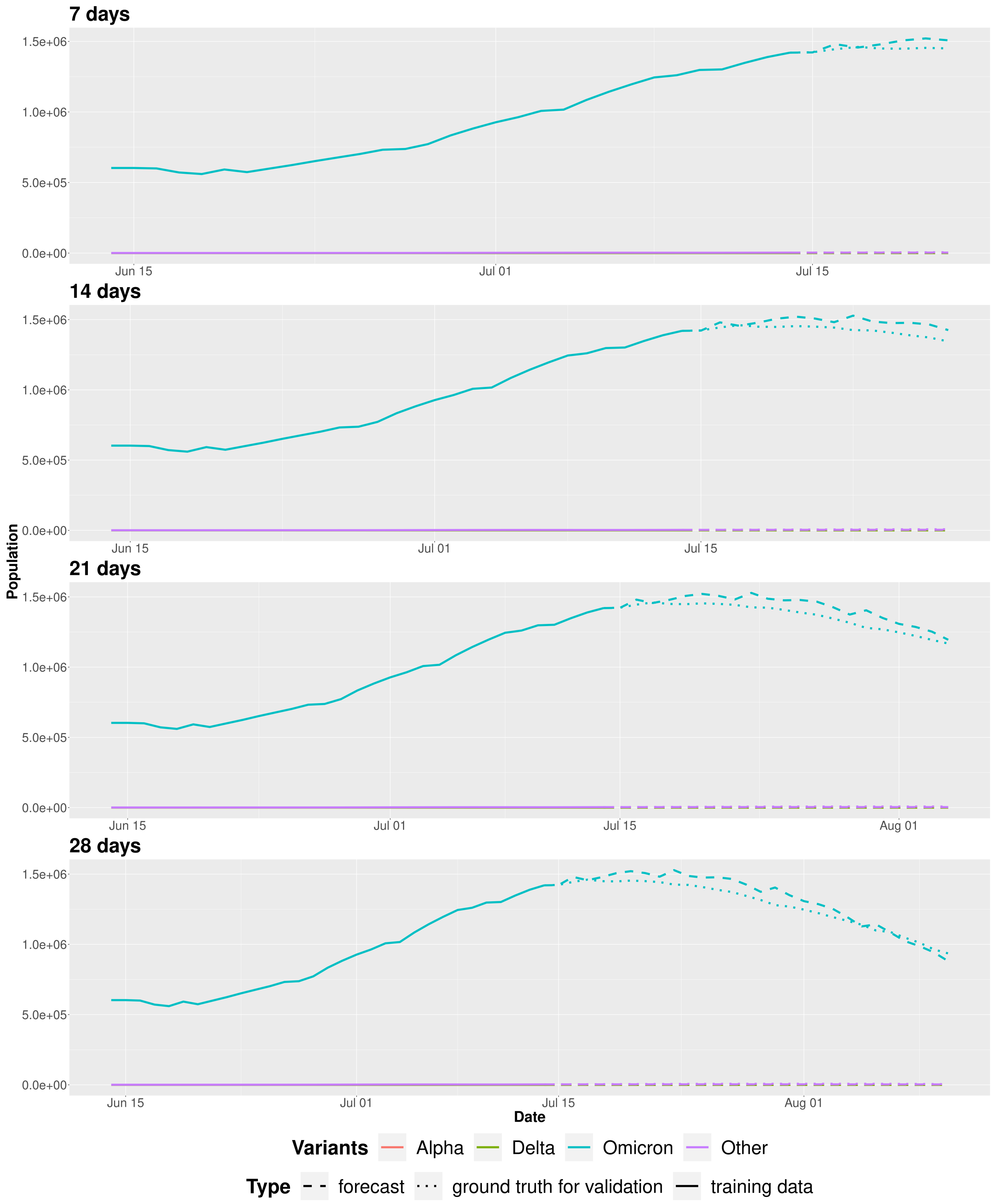


**Figure SM11.** Evolution of the $I_{Omicron}(\tilde{t})$ compartment using Sybil on the fourth scenario in which we forecast starting from July 14$^{th}$ 2022 (the dashed line shows the prediction, while solid and dotted lines represent the training data and the ground-truth values extracted from the surveillance data, respectively).

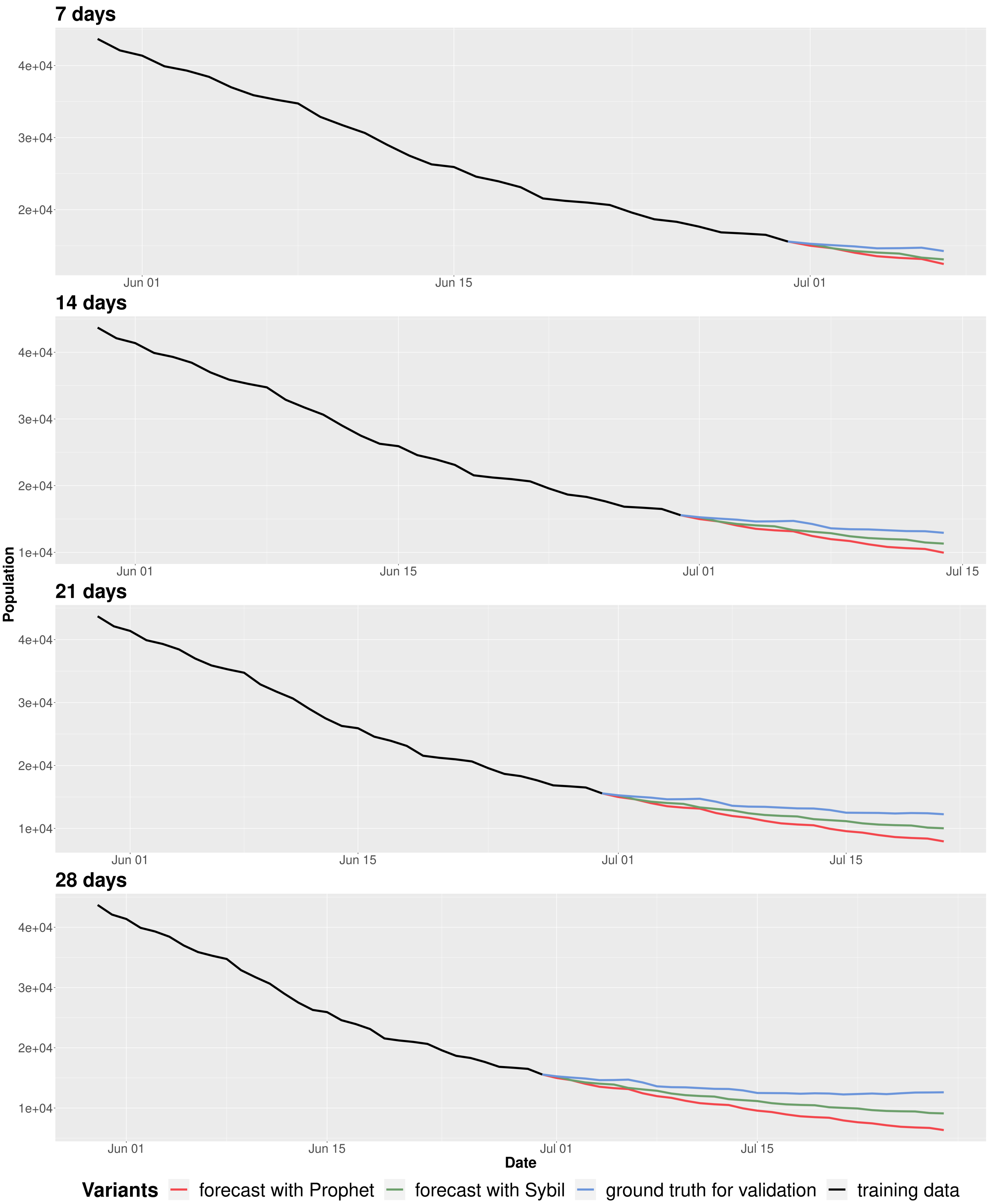


**Figure SM12.** The figure refer to the fifth scenario in which we forecast starting from June 30th 2020 and shows the evolution of the $I_{Other}(\tilde{t})$ compartment using Sybil (green line) and Prophet (red line) using the same period as training data (black line) comparing and contrasting the predictions against the surveillance data for the period spanning the forecasting window (blue line).

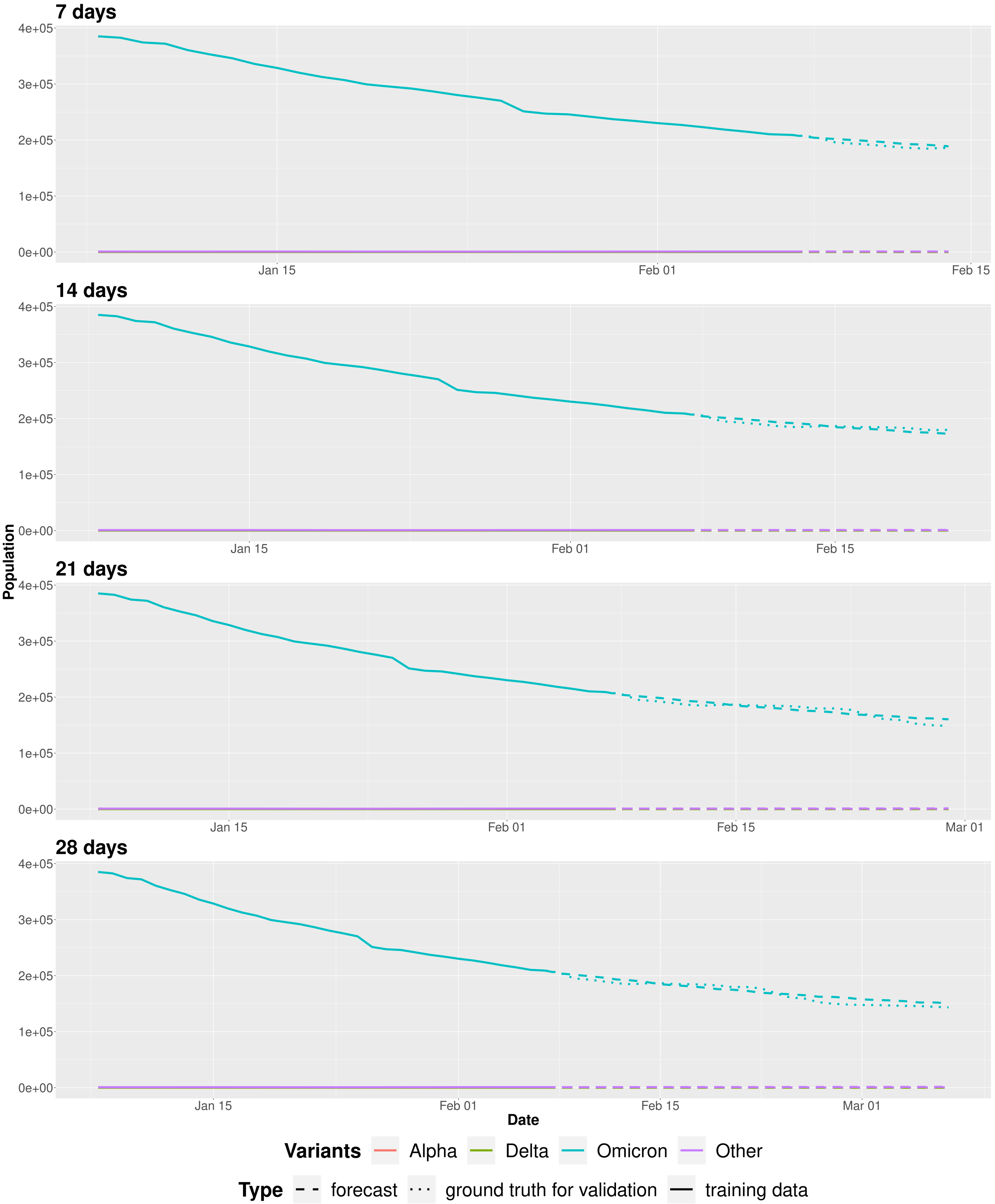


**Figure SM13.** Evolution of the $I_{Omicron}(\tilde{t})$ compartment using Sybil on the sixth scenario in which we forecast starting from February 7$^{th}$ 2023 (the dashed line shows the prediction, while solid and dotted lines represent the training data and the ground-truth values extracted from the surveillance data, respectively).

##### 4.1.1 Relative errors

Table SM1 shows the relative errors (mean, standard deviation, minimum and maximum values) between the ground truth used for validation and the obtained forecast with both approaches for all the considered scenarios in Italy. This table confirms that Sybil almost always performs better than Prophet. When there are small changes or no changes in the trajectory, both approaches works well (but also in these cases Sybil works slightly better). For the third scenario we do not report the errors for the two-, three- and four-weeks forecast because they are too high.

##### 4.1.2 Fixed recovery rate

Due to the limited availability of data on recoveries, we tried to use a fixed recovery rate $\frac{1}{\gamma}$ equal to 14 days – same for each variant. However, another possible extension could be to estimate a different recovery rate that depends on variants. The results obtained are very similar to the ones obtained with a time-dependent recovery rate. Figure SM14 shows the three scenario that we used to test Sybil with a fixed recovery rate – the three selected scenarios are the same of the first three in Figure SM6. Figures SM15, SM16 and SM17 show the predictions obtained in the three considered scenarios. From these figures we can see that the use of a fixed recovery rate does not alter the course of infection too much, at most it anticipates it or delays it by a few days.

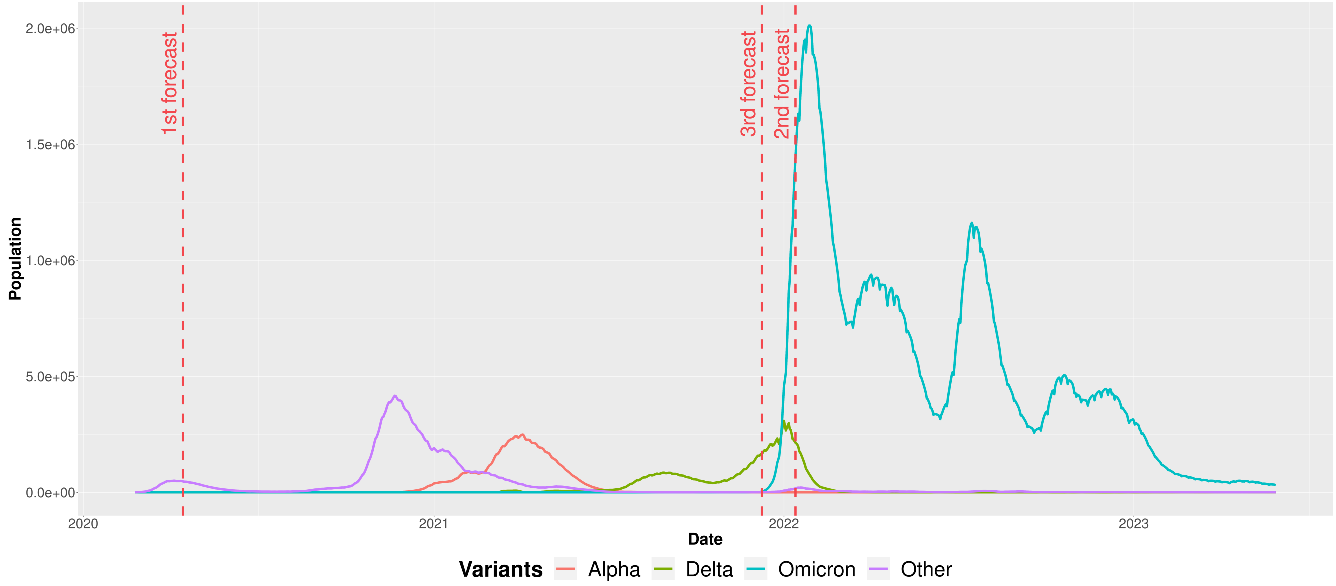


**Figure SM14.** Evolution of the $I_v(\tilde{t})$ compartments from February 2020 to May 2023 in Italy with the three considered forecast scenarios. Vertical dashed lines mark the selected dates to test Sybil's forecasting using a fixed recovery rate instead of a time-dependent one.

**Table SM1.** Mean relative errors with standard deviation and minimum and maximum values between the ground truth used for validation and the obtained forecast with Sybil and Prophet for the six considered scenarios in Italy.

| Scenario | Approach | 1 week | | | | 2 weeks | | | | 3 weeks | | | | 4 weeks | | | |
|---|---|---|---|---|---|---|---|---|---|---|---|---|---|---|---|---|---|
| | | *min* | *max* | *mean* | *std* | *min* | *max* | *mean* | *std* | *min* | *max* | *mean* | *std* | *min* | *max* | *mean* | *std* |
| 1st | Prophet | **1.90e-4** | 0.019 | 0.009 | 0.007 | **1.90e-4** | 0.055 | 0.024 | 0.019 | **1.90e-4** | 0.111 | 0.045 | 0.036 | **1.90e-4** | 0.235 | 0.082 | 0.073 |
| | Sybil | 4.71e-4 | **0.006** | **0.003** | **0.002** | 4.71e-4 | **0.016** | **0.008** | **0.006** | 4.71e-4 | **0.031** | **0.014** | **0.011** | 4.71e-4 | **0.079** | **0.027** | **0.026** |
| 2nd | Prophet | 0.018 | 0.219 | 0.098 | 0.073 | 0.018 | 0.924 | 0.348 | 0.304 | 0.018 | 2.590 | 0.799 | 0.760 | 0.018 | 7.068 | 1.833 | 2.048 |
| | Sybil | **0.008** | **0.072** | **0.035** | **0.023** | **0.008** | **0.227** | **0.112** | **0.087** | **0.008** | **0.332** | **0.170** | **0.112** | **0.008** | **0.436** | **0.222** | **0.134** |
| 3rd (Dec 18th, 2021) | Prophet | **0.046** | **0.232** | **0.152** | **0.066** | - | - | - | - | - | - | - | - | - | - | - | - |
| | Sybil | 0.055 | 0.675 | 0.403 | 0.224 | - | - | - | - | - | - | - | - | - | - | - | - |
| 3rd (Dec 27th, 2021) | Prophet | 0.029 | 0.150 | 0.104 | **0.047** | - | - | - | - | - | - | - | - | - | - | - | - |
| | Sybil | **0.014** | **0.135** | **0.081** | 0.049 | - | - | - | - | - | - | - | - | - | - | - | - |
| 4th | Prophet | 0.054 | 0.147 | 0.105 | 0.033 | 0.054 | 0.251 | 0.158 | 0.063 | 0.054 | 0.377 | 0.214 | 0.097 | 0.054 | 0.528 | 0.279 | 0.143 |
| | Sybil | **0.023** | **0.100** | **0.065** | **0.027** | **0.023** | **0.137** | **0.096** | **0.038** | **0.023** | **0.148** | **0.111** | **0.037** | **0.023** | **0.148** | **0.115** | **0.034** |
| 5th | Prophet | 0.004 | 0.032 | 0.018 | 0.010 | 0.004 | 0.058 | 0.031 | 0.017 | 0.004 | 0.088 | 0.045 | 0.025 | 0.004 | 0.124 | 0.061 | 0.036 |
| | Sybil | **2.63e-4** | **0.023** | **0.012** | **0.008** | **2.63e-4** | **0.032** | **0.018** | **0.009** | **2.63e-4** | **0.046** | **0.025** | **0.013** | **2.63e-4** | **0.069** | **0.034** | **0.019** |
| 6th | Prophet | 0.007 | **0.064** | **0.040** | **0.019** | 0.007 | **0.348** | **0.134** | **0.118** | 0.007 | **0.407** | **0.218** | **0.155** | 0.007 | **0.407** | **0.234** | **0.138** |
| | Sybil | **0.004** | 0.080 | 0.042 | 0.026 | **0.004** | 0.404 | 0.147 | 0.134 | **0.004** | 0.518 | 0.259 | 0.196 | **0.004** | 0.518 | 0.298 | 0.183 |

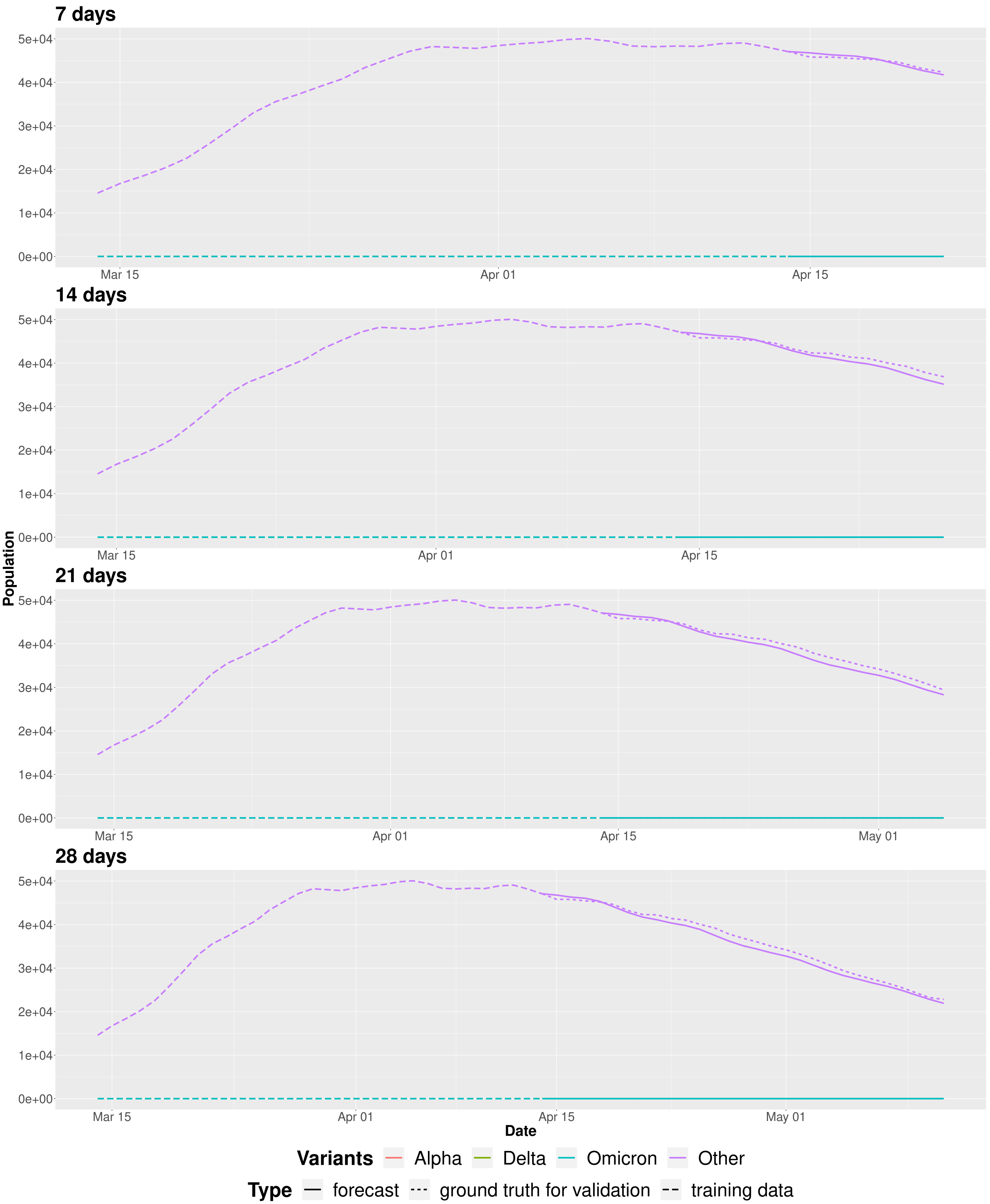


**Figure SM15.** Evolution of the $I_{Other}(\tilde{t})$ compartment using Sybil with a fixed recovery rate on the first scenario in which we forecast starting from April 14$^{\text{th}}$ 2020 (the dashed line shows the prediction, while solid and dotted lines represent the training data and the ground-truth values extracted from the surveillance data, respectively).

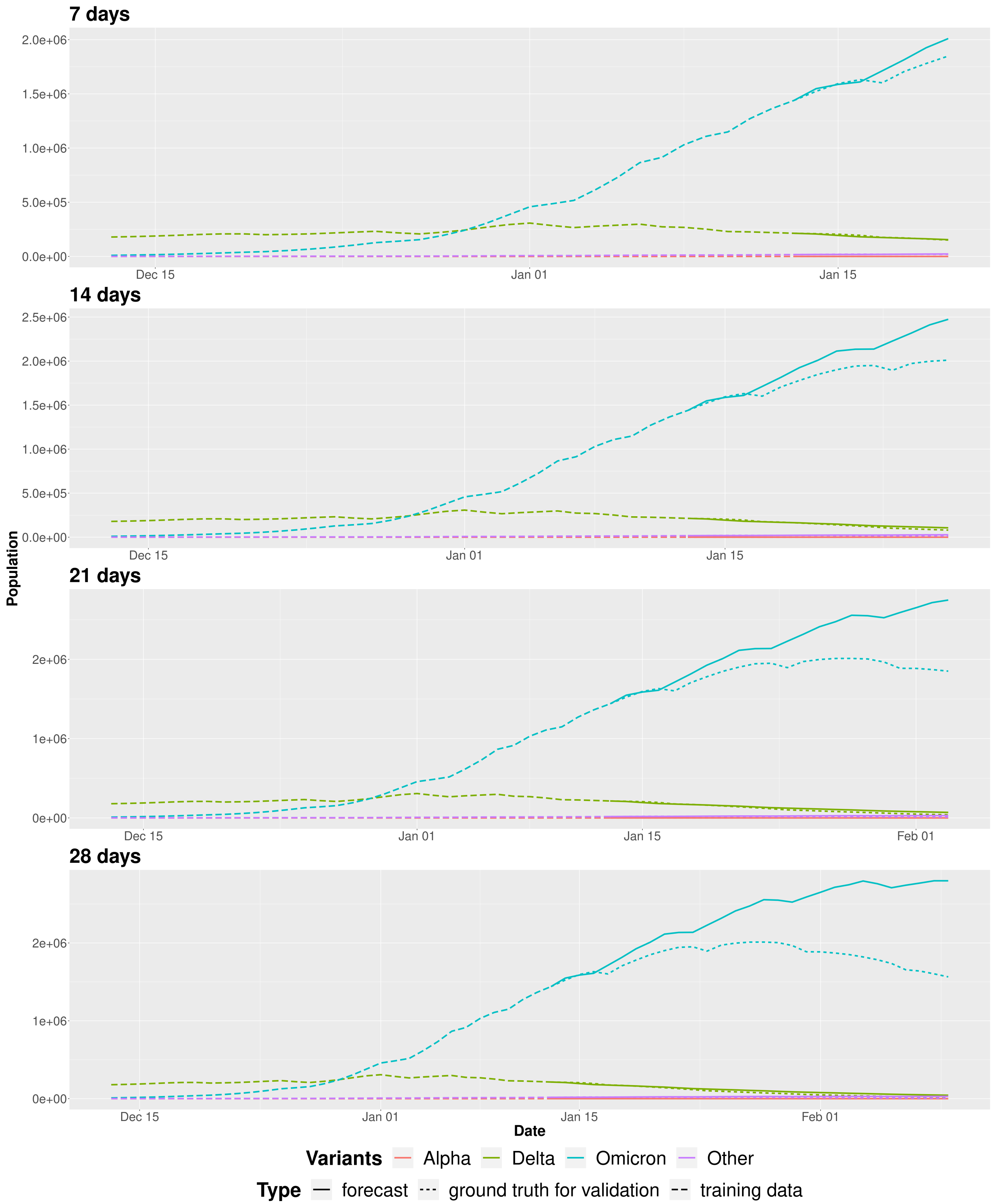


**Figure SM16.** Evolution of the $I_v(\tilde{t})$ compartments using Sybil with a fixed recovery rate on the second scenario in which we forecast starting from January 13$^{\text{th}}$ 2022 (the dashed line shows the prediction, while solid and dotted lines represent the training data and the ground-truth values extracted from the surveillance data, respectively).

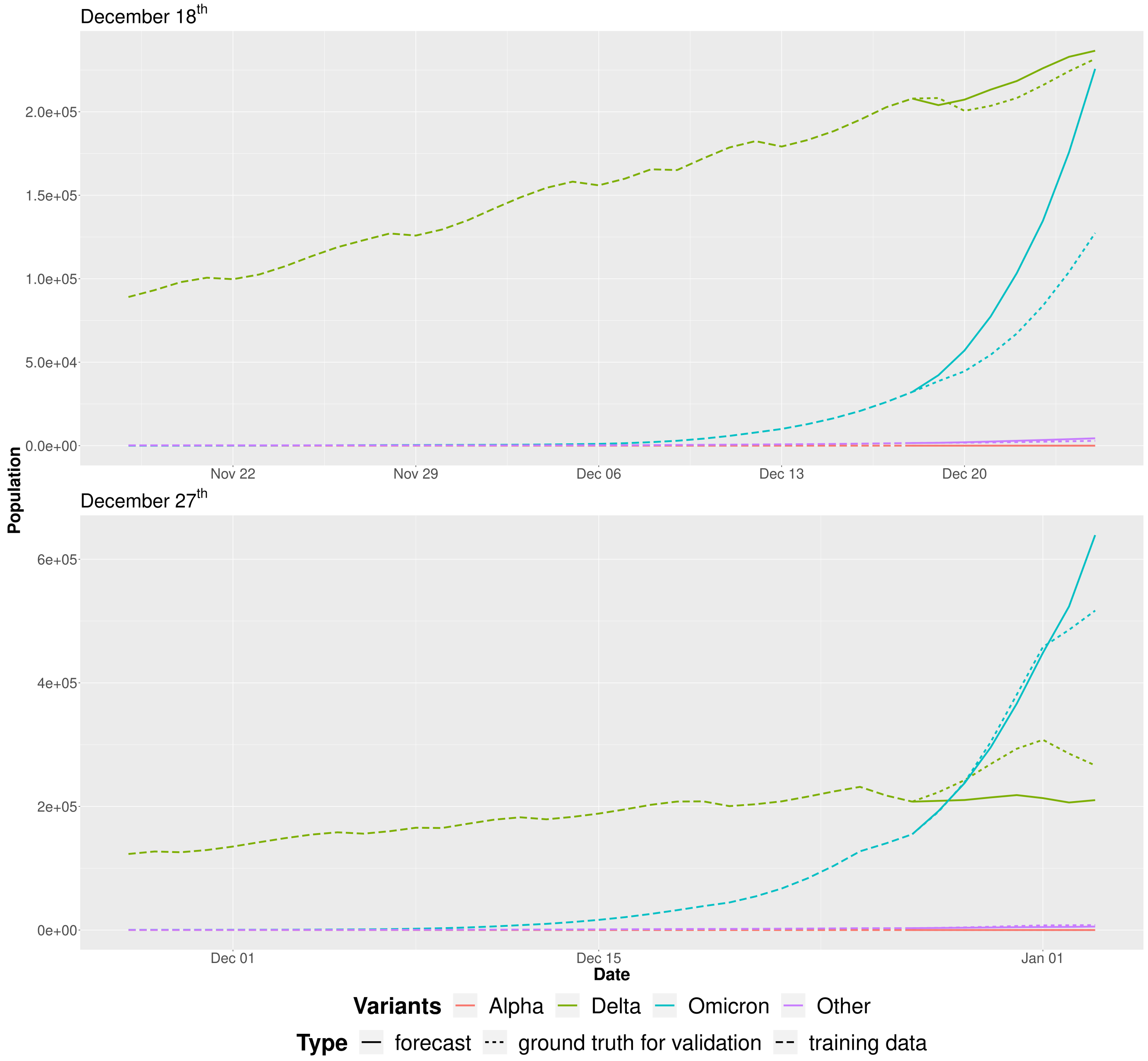


**Figure SM17.** Evolution of the $I_v(\tilde{t})$ compartments using Sybil on the third scenario with a fixed recovery rate. In the first plot we forecast starting from December 18$^{th}$, 2021, while in second one we forecast starting from December 27$^{th}$, 2021. Both the plots refer to a forecast one week into the future. The dashed line shows the prediction, while solid and dotted lines represent the training data and the ground-truth values extracted from the surveillance data, respectively.

#### 4.2 Results with V $= 11$

We tried to use a different aggregation of lineages (V $= 11$). Figures SM18, SM19 and SM20 show the proportion of each variant, the evolution of the $I_v(\tilde{t})$ compartments and the evolution of the infection rates for each variant $v$ from February 2020 to May 2023 in Italy, respectively. With this configuration, we considered again the main peak of infections. Figure SM21 shows how the forecast changes moving the training window from January 10$^{th}$, 2022 to January 15$^{th}$, 2022 by one day and how Sybil can capture the change of slope of the BA.1 variant – Figure SM22 shows how Sybil is able to capture the infection rates. We can also see a new emerging variant (BA.2). In particular, the first row of Figure SM21 shows that the model initially sees this new variant, while the second row shows that it would appear that we can't follow the trend. However, Figure SM23 shows that we have obtained good predictions on the infection rates of the BA.2 variant. This behaviour is due to the fact that, when there is a new emerging variant – or, more generally, in periods in which we have a trajectory which is initially growing –, we have small values for the rates and Sybil sees an exponential growth. Indeed, from a quantitative point of view, we may not be able to follow precisely the trajectory, but from a qualitative point of view we are able to capture the new emerging variant/outbreak. Figure SM24 shows that after one week we capture well the trajectory of the same variant (especially after one and four weeks). We point out that the model will only see a new emerging variant if it is already present in the training data.

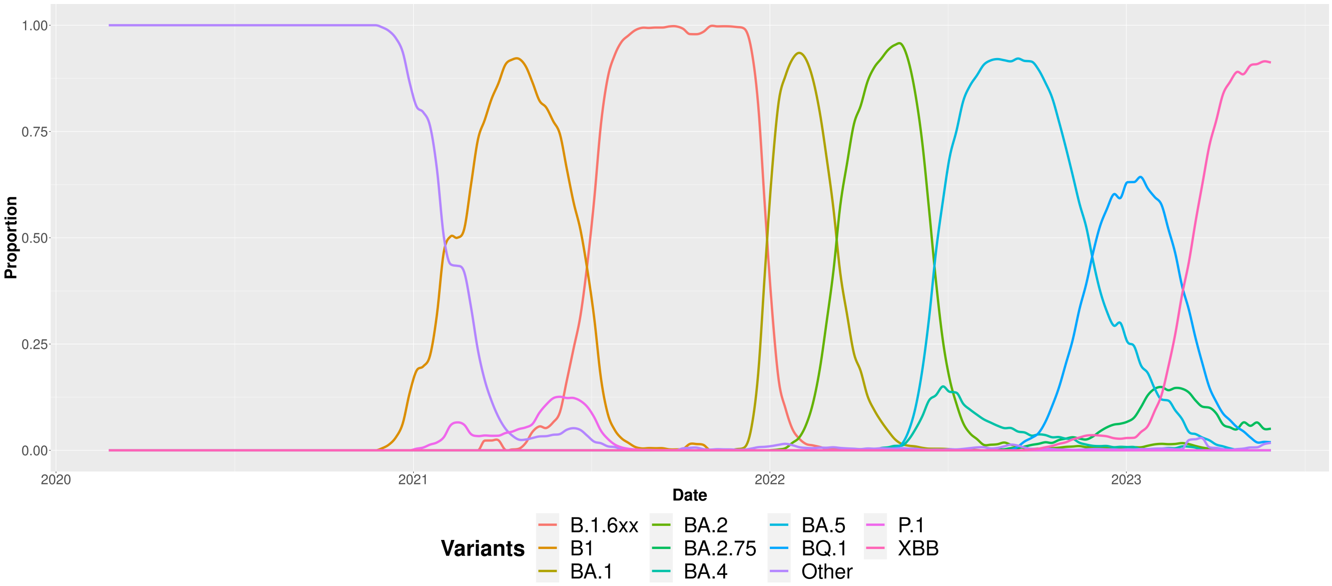


**Figure SM18.** Proportion of the aggregated variants from February 2020 to May 2023 in Italy.

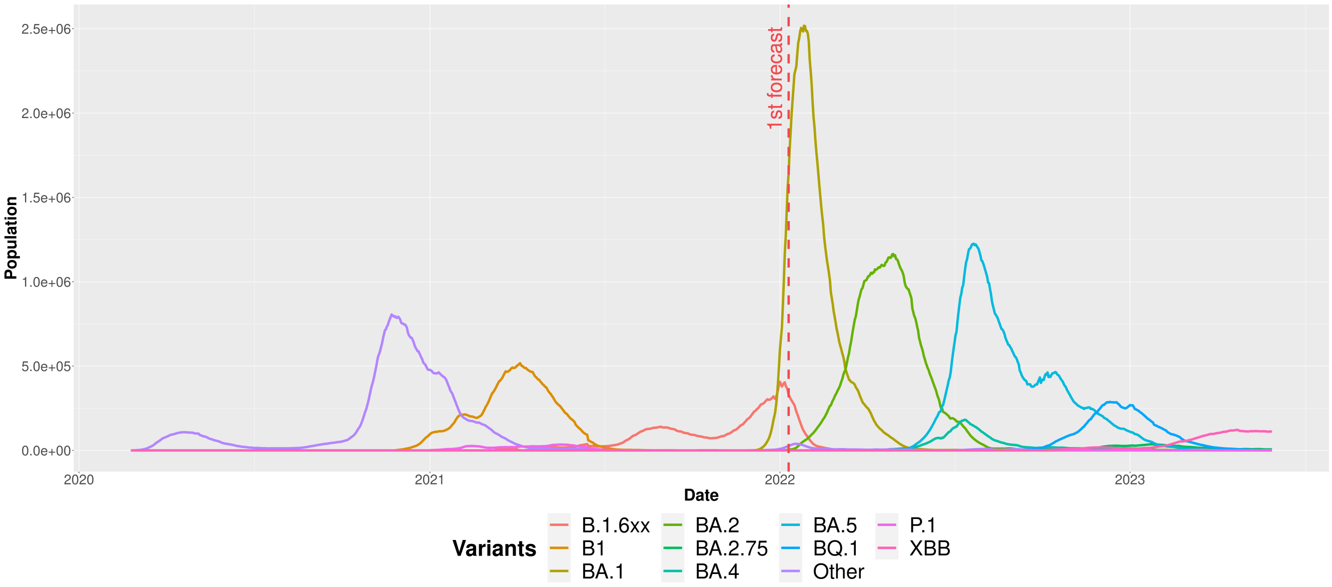


**Figure SM19.** Evolution of the $I_v(\tilde{t})$ compartments from February 2020 to May 2023 in Italy with the selected forecast scenario.

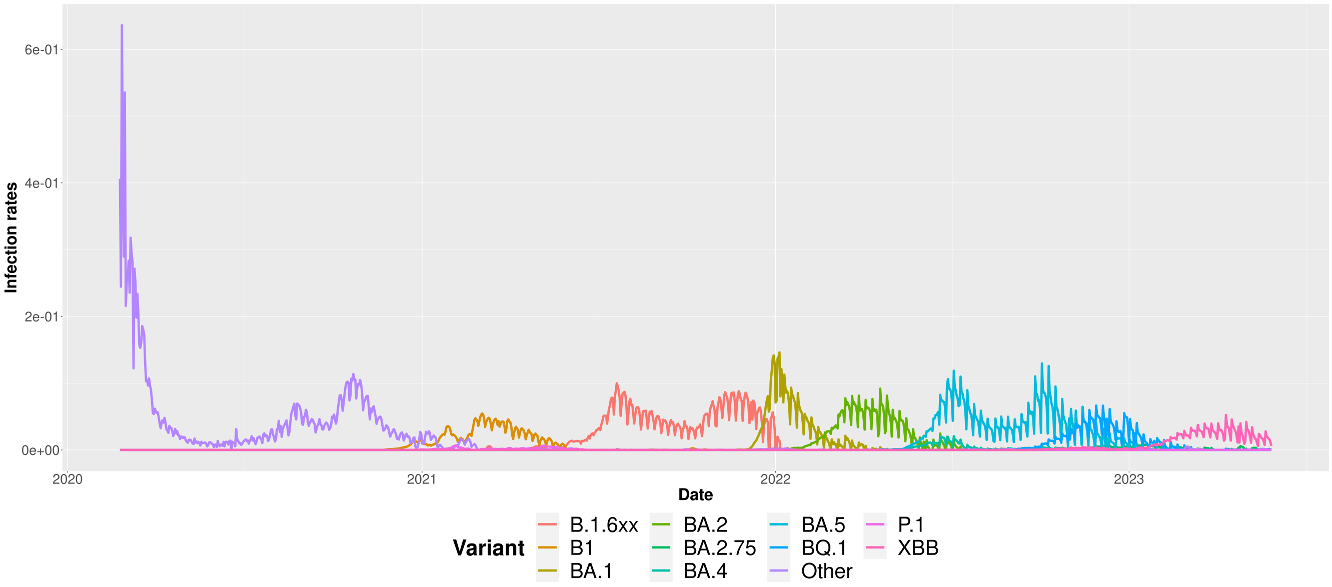


**Figure SM20.** Evolution of the infection rates $\beta_v(\tilde{t})$ for each variant from February 2020 to May 2023 in Italy.

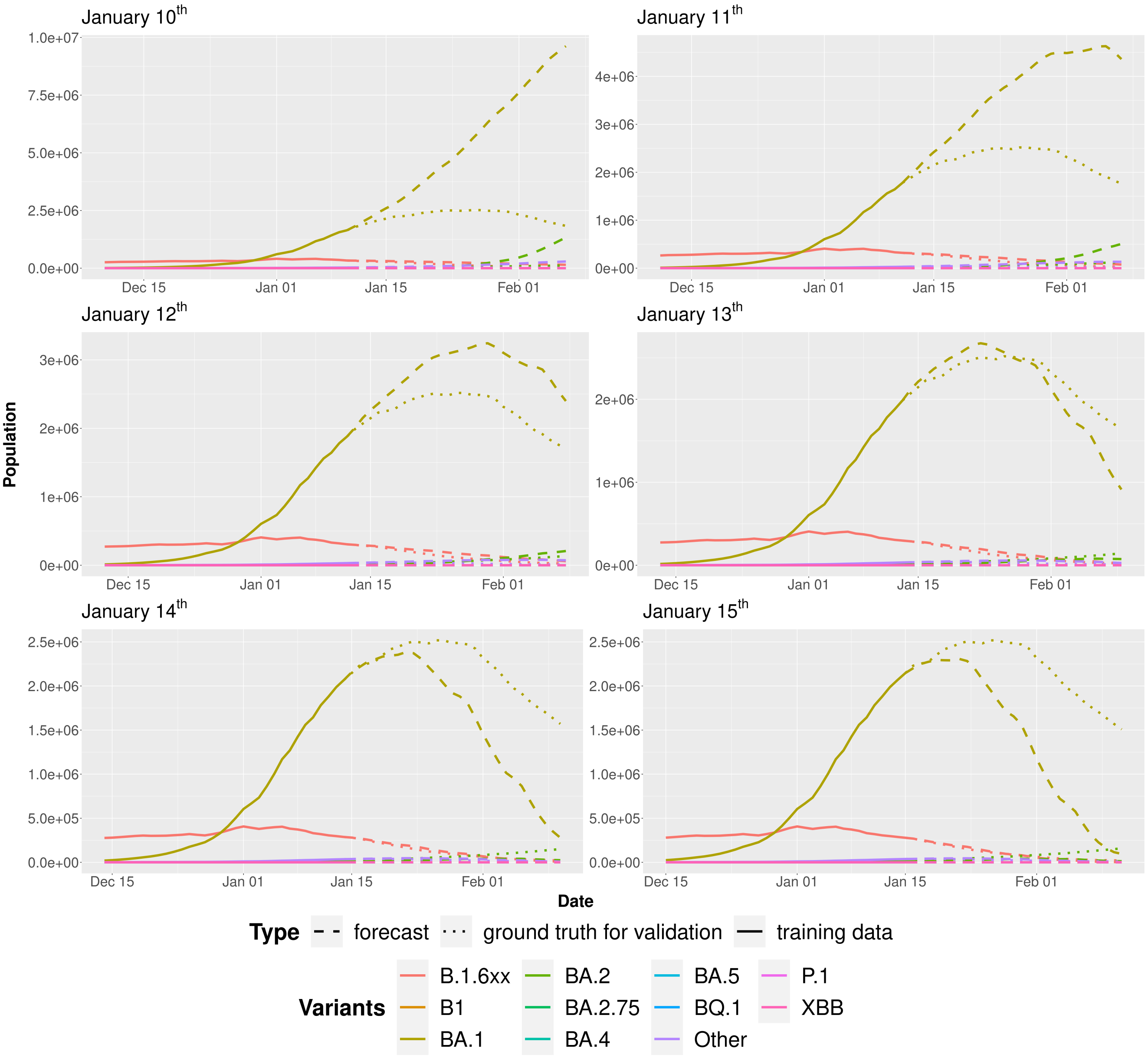


**Figure SM21.** Evolution of the $I_v(\tilde{t})$ compartments using Sybil starting the forecast from January 10$^{th}$, 11$^{th}$, 12$^{th}$, 13$^{th}$, 14$^{th}$ and 15$^{th}$, 2022 and moving the training window by one day (the dashed line shows the prediction, while solid and dotted lines represent the training data and the ground-truth values extracted from the surveillance data, respectively). All plots refer to a forecast four weeks into the future.

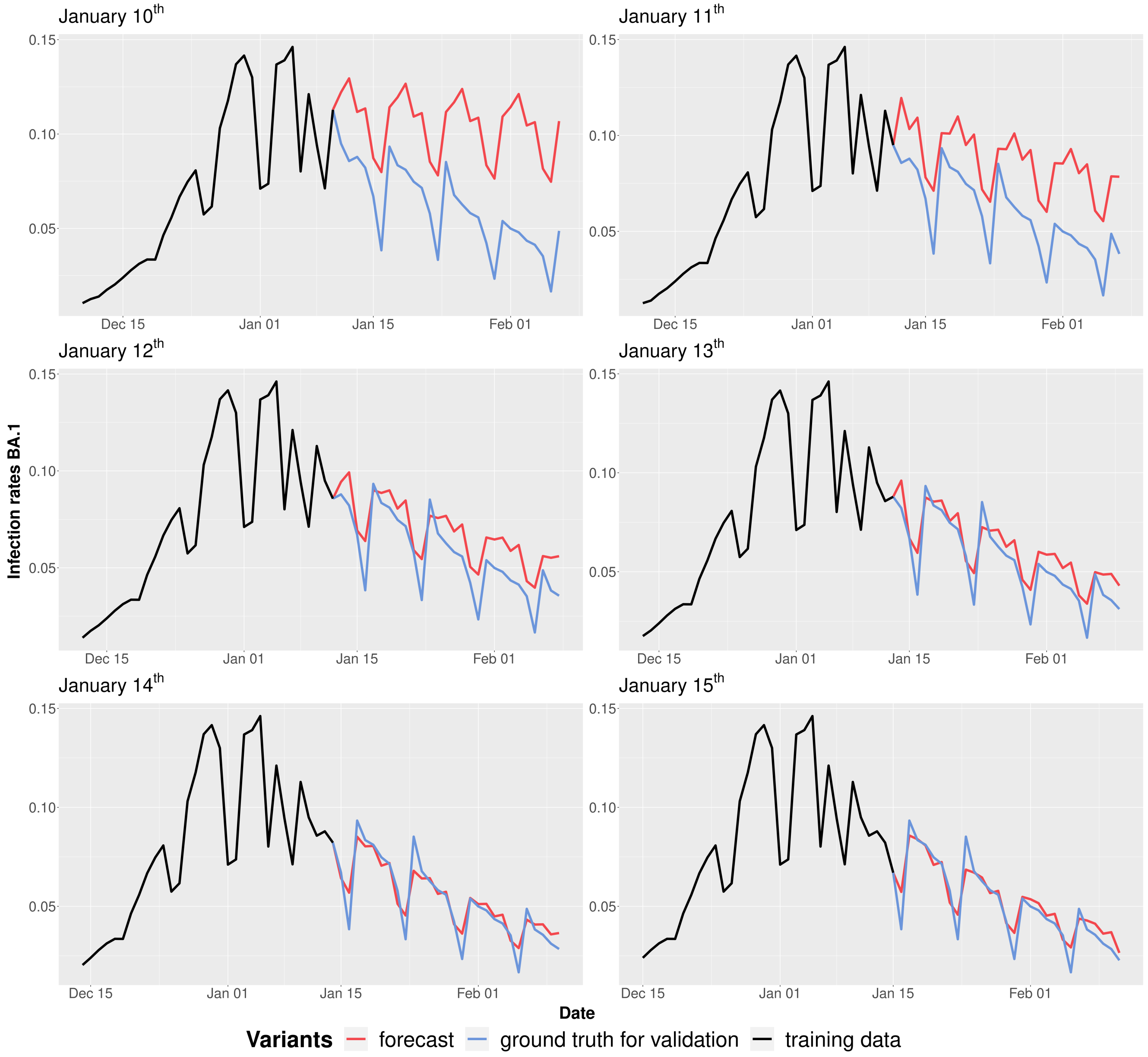


**Figure SM22.** Evolution of the infection rates $\beta_{BA.1}(\tilde{t})$ starting the forecast from January $10^{th}$, $11^{th}$, $12^{th}$, $13^{th}$, $14^{th}$ and $15^{th}$, 2022 and moving the training window by one day (the red line shows the prediction, while black and blue lines represent the training data and the ground-truth values extracted from the surveillance data, respectively). All plots refer to a forecast four weeks into the future.

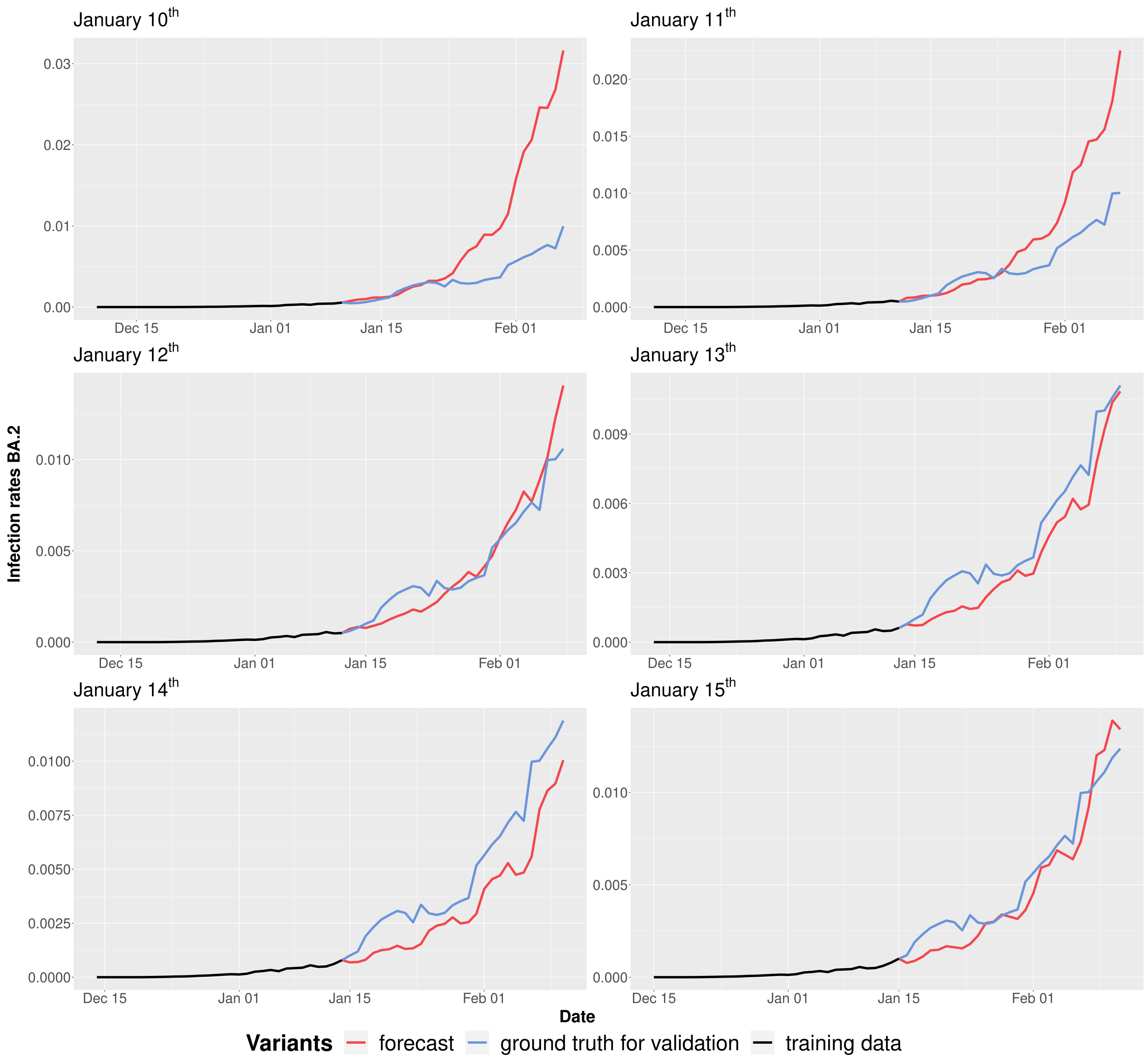


**Figure SM23.** Evolution of the infection rates $\beta_{BA.2}(\tilde{t})$ starting the forecast from January 10$^{th}$, 11$^{th}$, 12$^{th}$, 13$^{th}$, 14$^{th}$ and 15$^{th}$, 2022 moving the training window by one day (the red line shows the prediction, while black and blue lines represent the training data and the ground-truth values extracted from the surveillance data, respectively). All plots refer to a forecast four weeks into the future.

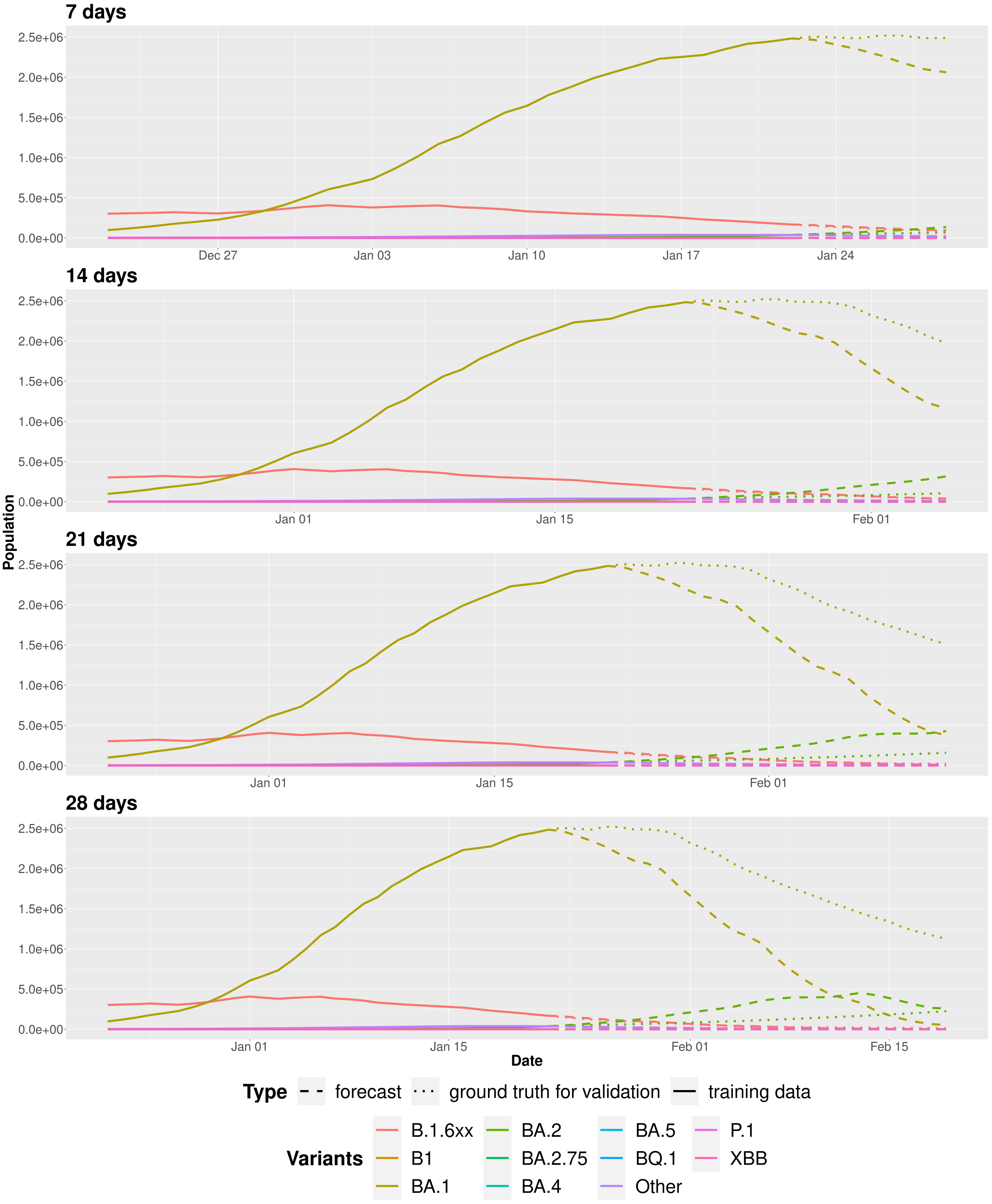


**Figure SM24.** Evolution of the $I_v(\tilde{t})$ compartments using Sybil starting the forecast from January 22$^{\text{th}}$, 2022 (the dashed line shows the prediction, while solid and dotted lines represent the training data and the ground-truth values extracted from the surveillance data, respectively).

### Other results on Austria

Figure SM25 shows the evolution of the $I(\tilde{t})$ compartment from February 2020 to May 2023 in Austria. Figures SM26, SM27 and SM28 show the evolution of infection, recovery and fatality rates respectively, in the same period. Figures SM29, SM30 and SM31 show the daily proportion of each variant, the evolution of the $I_v(\tilde{t})$ compartments and the evolution of infection rates for each variant in the same period, respectively. Figure SM30 shows the previously considered forecast scenario (first) and other three new scenarios (second, third and fourth). Figures SM32, SM33 and SM34 show the results obtained with the second, third and fourth scenarios.

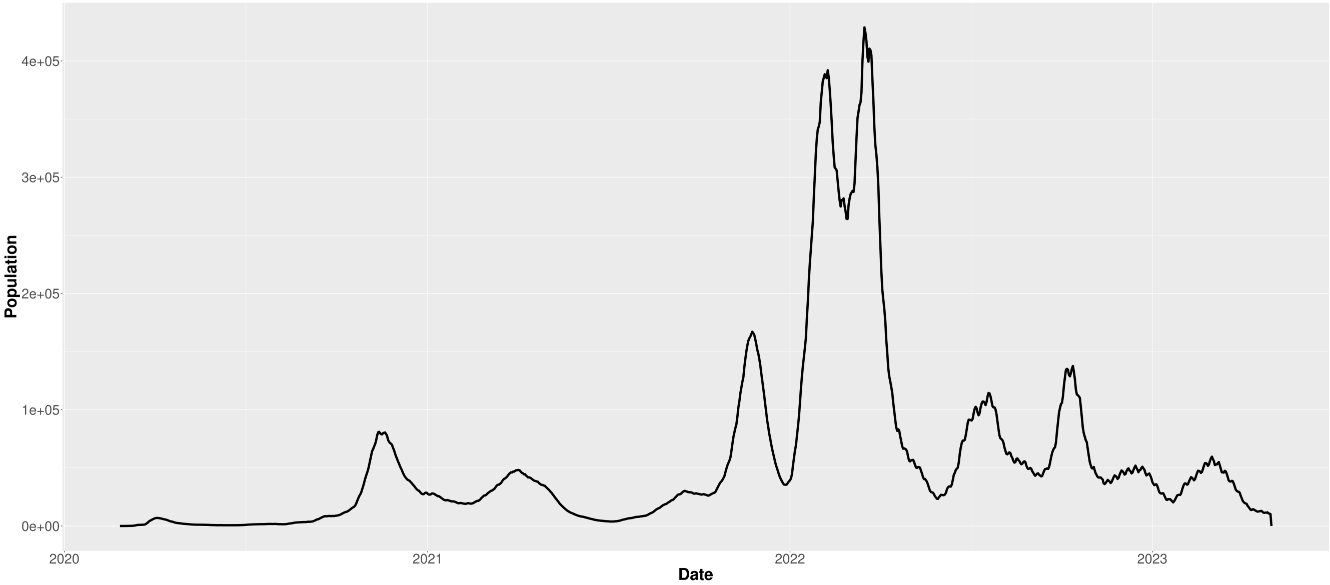


**Figure SM25.** Evolution of the $I(\tilde{t})$ compartment from February 2020 to May 2023 in Austria.

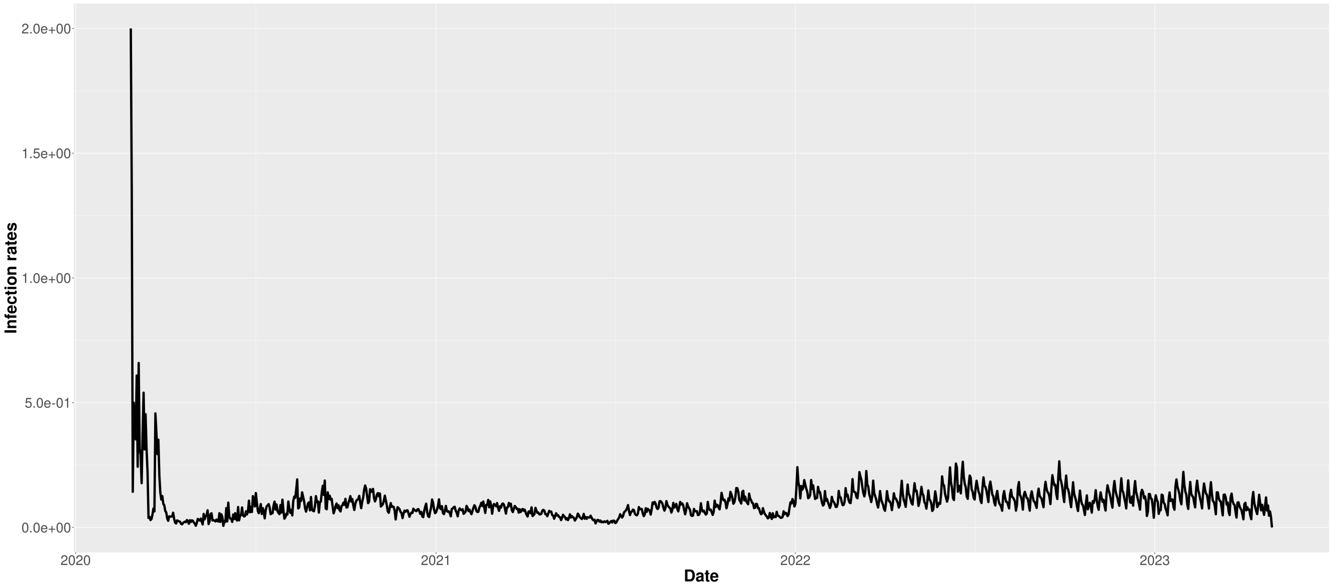


**Figure SM26.** Evolution of the infection rates $\beta(\tilde{t})$ from February 2020 to May 2023 in Austria.

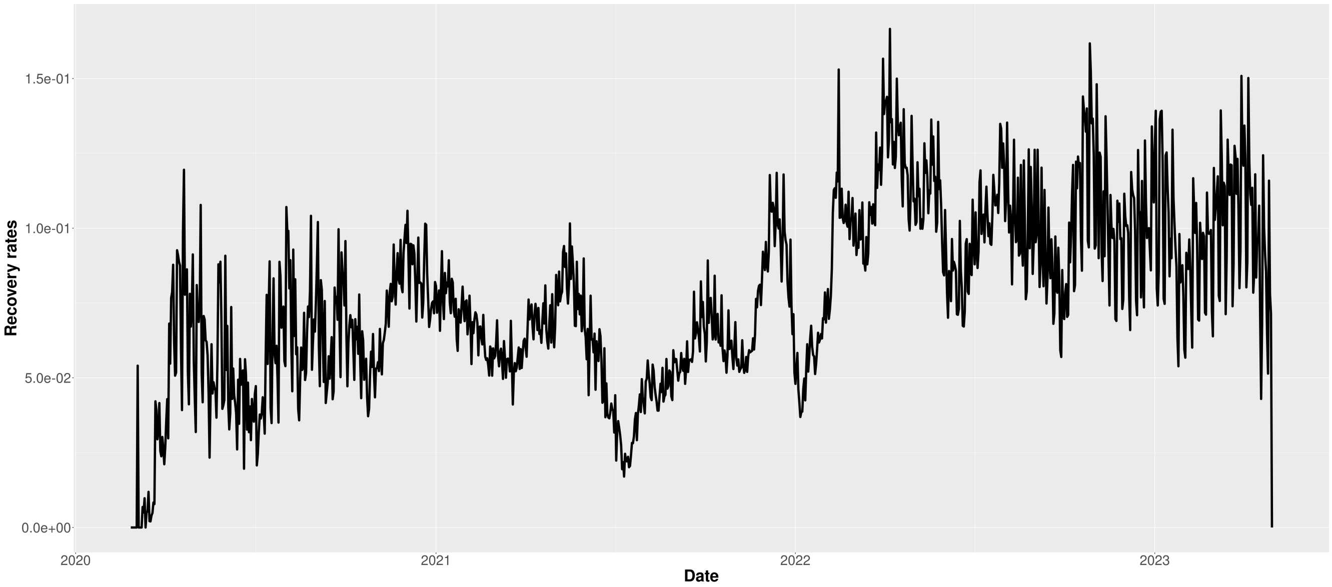


**Figure SM27.** Evolution of the recovery rates $\gamma(\tilde{t})$ from February 2020 to May 2023 in Austria.

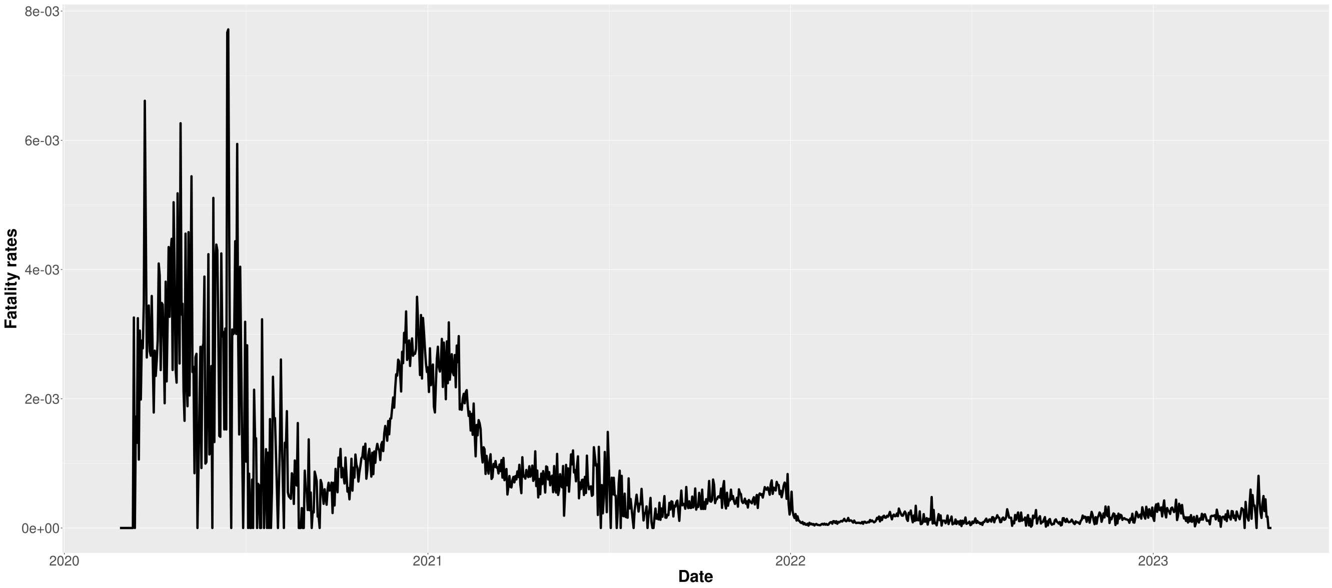


**Figure SM28.** Evolution of the fatality rates $\lambda(\tilde{t})$ from February 2020 to May 2023 in Austria.

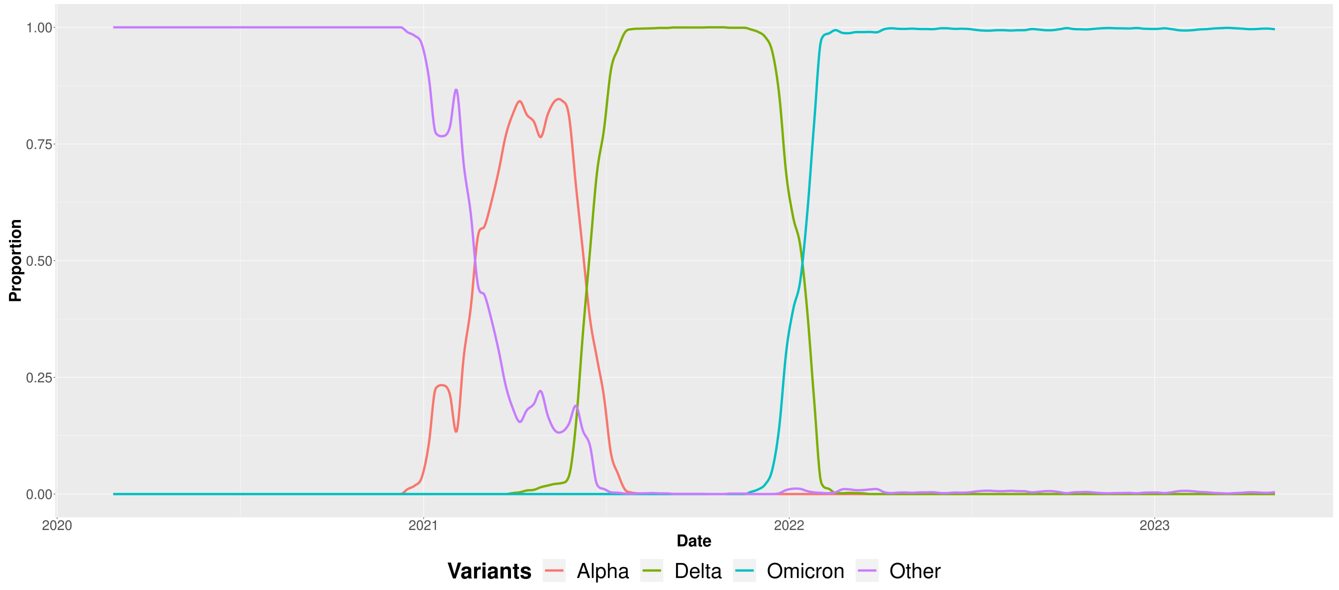


**Figure SM29.** Proportion of the aggregated variants from February 2020 to May 2023.

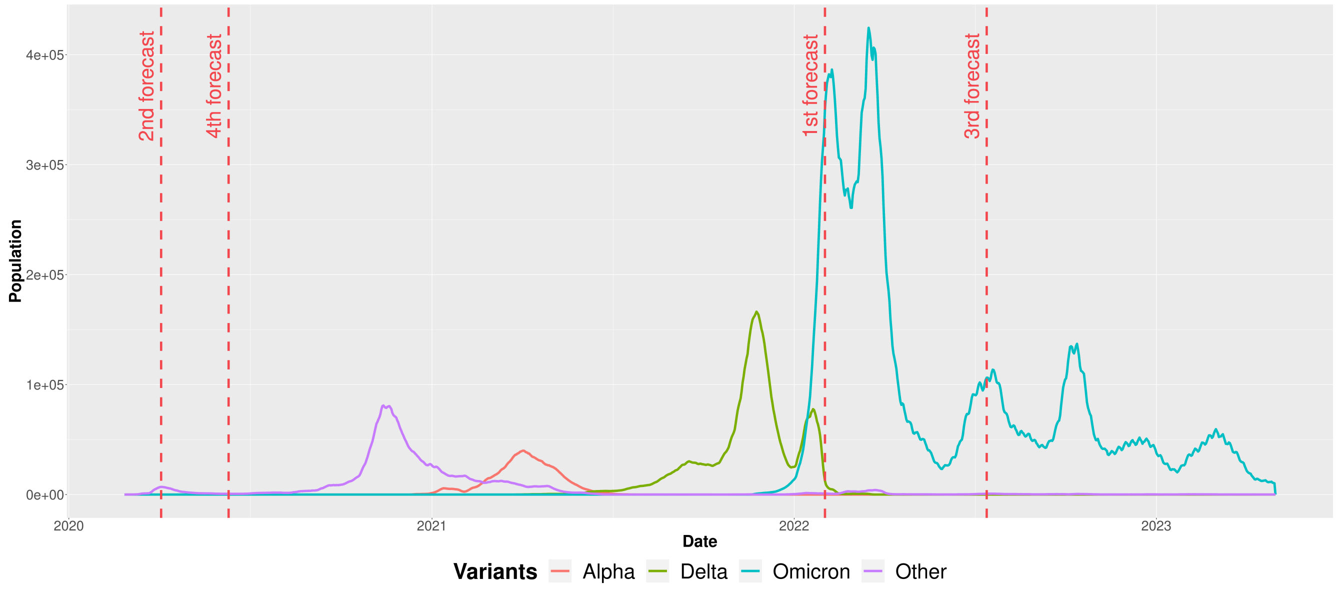


**Figure SM30.** Evolution of the $I_v(\tilde{t})$ compartments from February 2020 to May 2023 in Austria with the four considered forecast scenarios.

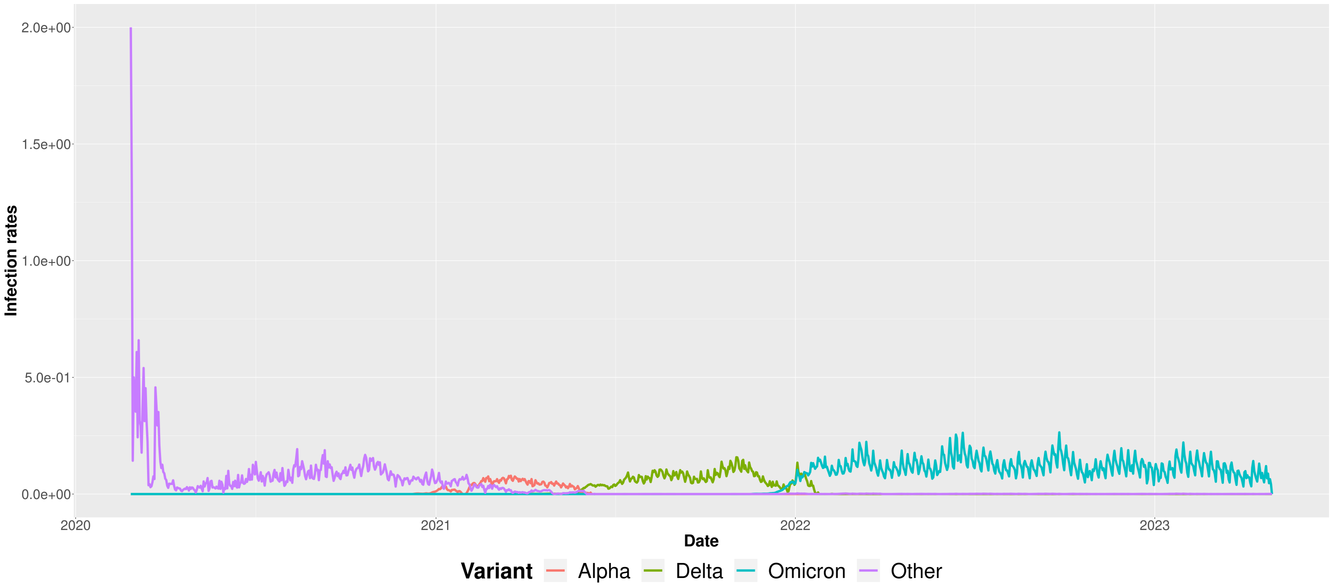


**Figure SM31.** Evolution of the infection rates $\beta_v(\tilde{t})$ for each variant from February 2020 to May 2023 in Austria.

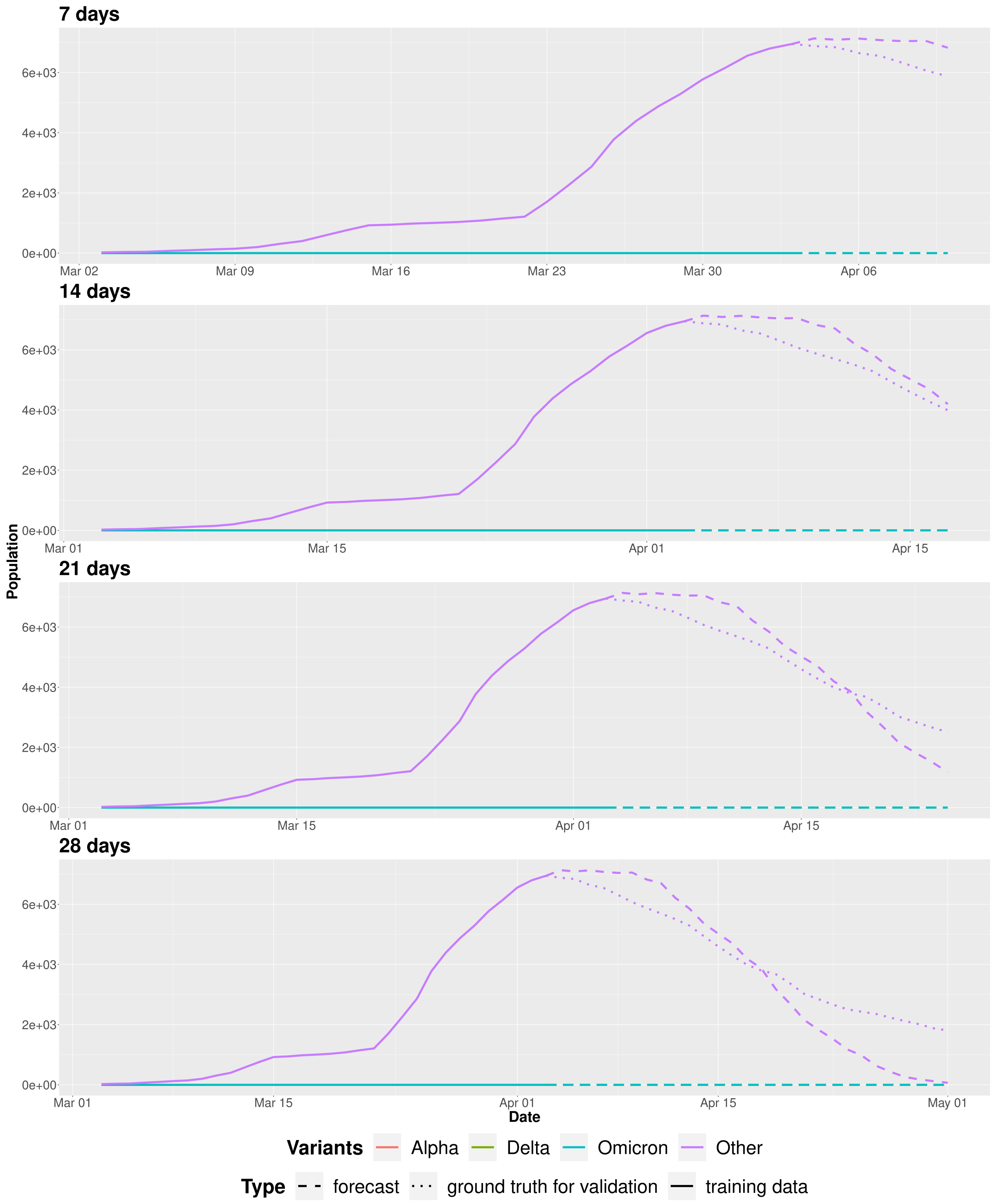


**Figure SM32.** Evolution of the $I_{Other}(\tilde{t})$ compartment using Sybil on the second scenario in which we forecast starting from April 3rd 2020 (the dashed line shows the prediction, while solid and dotted lines represent the training data and the ground-truth values extracted from the surveillance data, respectively).

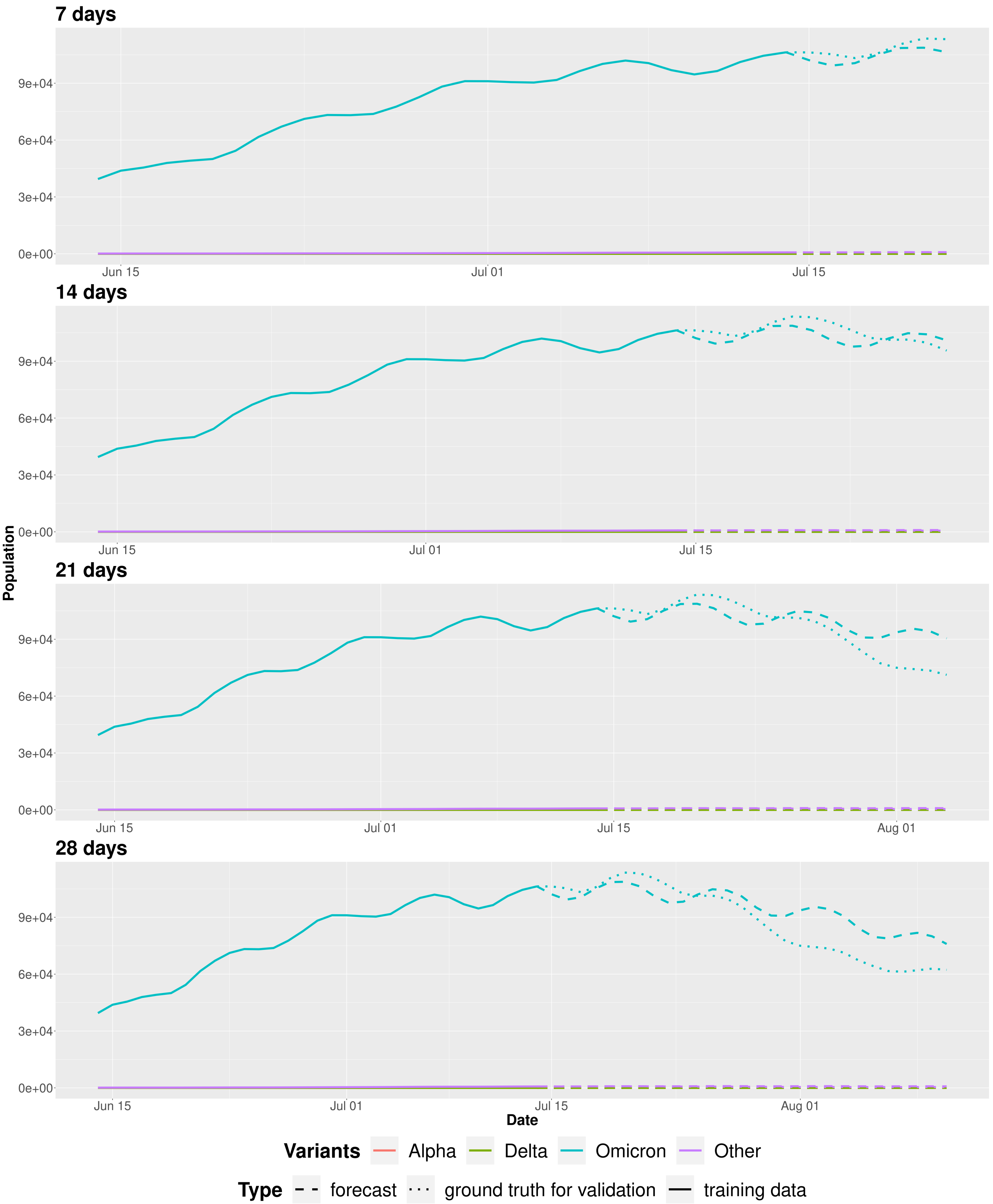


**Figure SM33.** Evolution of the $I_{Omicron}(\tilde{t})$ compartment using Sybil on the third scenario in which we forecast starting from July 14$^{th}$ 2022 (the dashed line shows the prediction, while solid and dotted lines represent the training data and the ground-truth values extracted from the surveillance data, respectively).

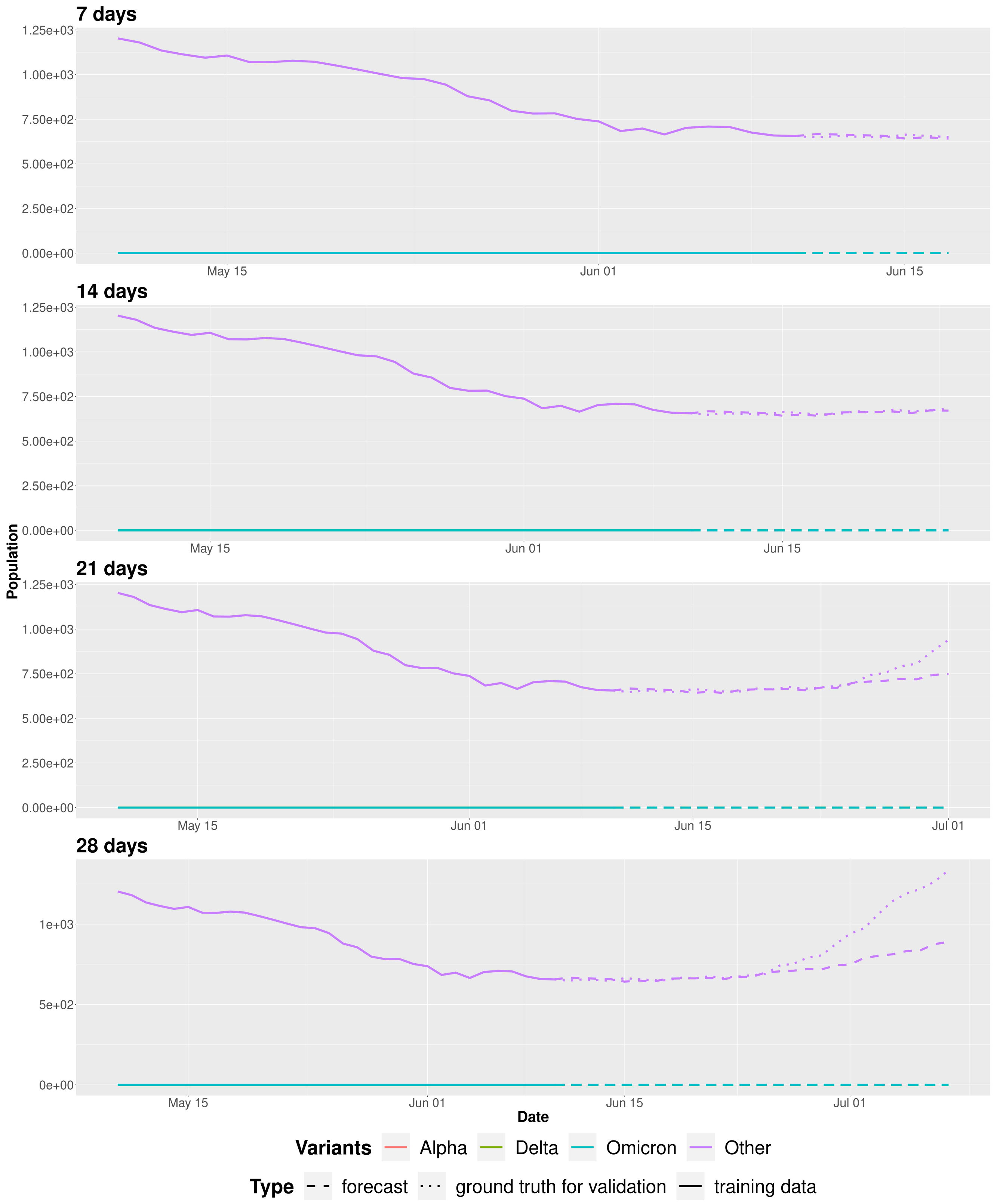


**Figure SM34.** Evolution of the $I_{Other}(\tilde{t})$ compartment using Sybil on the fourth scenario in which we forecast starting from June 10[th] 2020 (the dashed line shows the prediction, while solid and dotted lines represent the training data and the ground-truth values extracted from the surveillance data, respectively).

#### 5 Continuous monitoring

To set up a continuous monitoring system we have to obtain good predictions also in periods in which there is a new emerging variant or a new exploding outbreak. In the main paper we showed an example in which we forecast just before the exploding of the largest outbreak in Italy and, in a previous section, we showed another example using 11 different aggregation of lineages. In particular, considering again the case with 4 aggregation of lineages, Figure SM35 shows two scenarios in which we forecast just before a new outbreak – of the new emerging Omicron variant (first scenario, partly showed in the main paper) and of the Alpha variant (second scenario), respectively.

Figures SM36 and SM37 show an extension of the first ascending scenario presented in the main paper. In particular, these figures show how the one-week forecast changes moving the training window from December 9th, 2021 to January 11th, 2022 by three days. As already mentioned in the main document, in this case the upward trajectory is very steep, and Sybil finds it harder to calibrate with respect to the previous cases. Here, the Omicron variant is a new emerging variant and Sybil initially foresees a more aggressive exponential growth. Looking at Figure SM36 we can state that Sybil nailed the qualitative prediction, but needed more data to calibrate it. We can also see that Sybil captures well the prediction on the other active variant—the Delta variant.

In the second ascending scenario there are three active variants: Alpha, Delta and the *Other* variant. Figure SM38 shows how the one-week forecast changes moving the training window from February 15th, 2021 to March 20th, 2021 by three days. In particular, the Alpha variant is ascending while the *Other* variant is descending. The first three rows show that Sybil initially sees an exponential growth for the Alpha variant – see also Figure SM39 for the predictions on the infection rates. This exponential growth increases as the forecasted period increases. Again, we can state that Sybil nailed the qualitative prediction, but needs more data to calibrate the quantitative prediction – see the last two rows of Figure SM38.

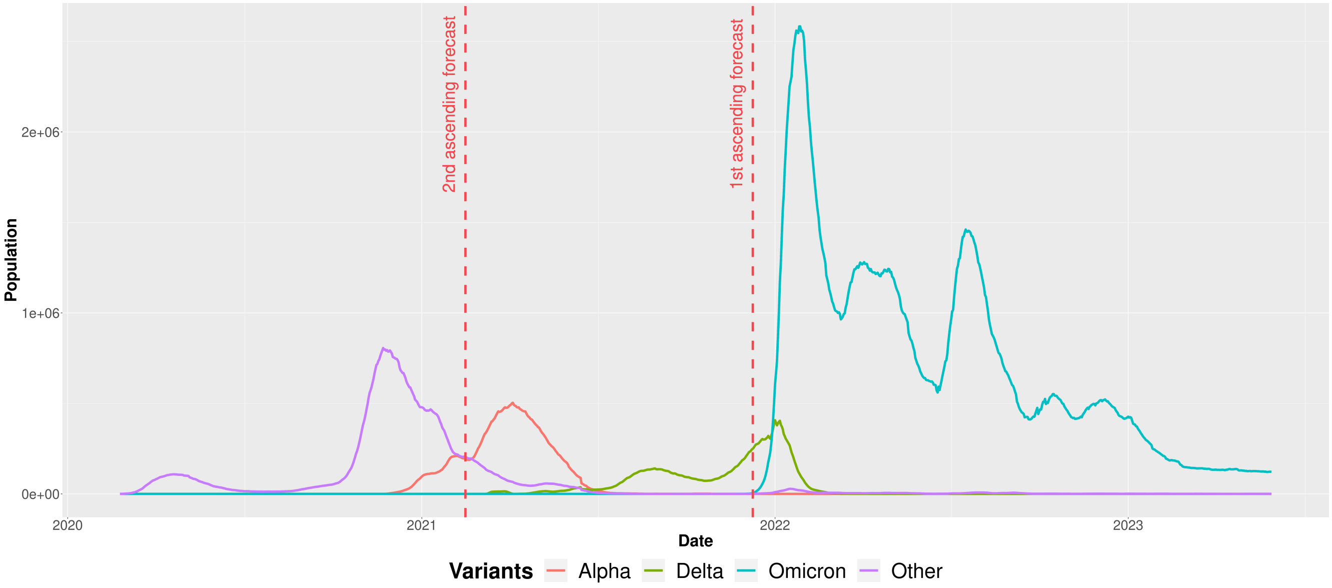


**Figure SM35.** Evolution of the $I_v(\tilde{t})$ compartment from February 2020 to May 2023 in Italy with the two considered ascending forecast scenarios.

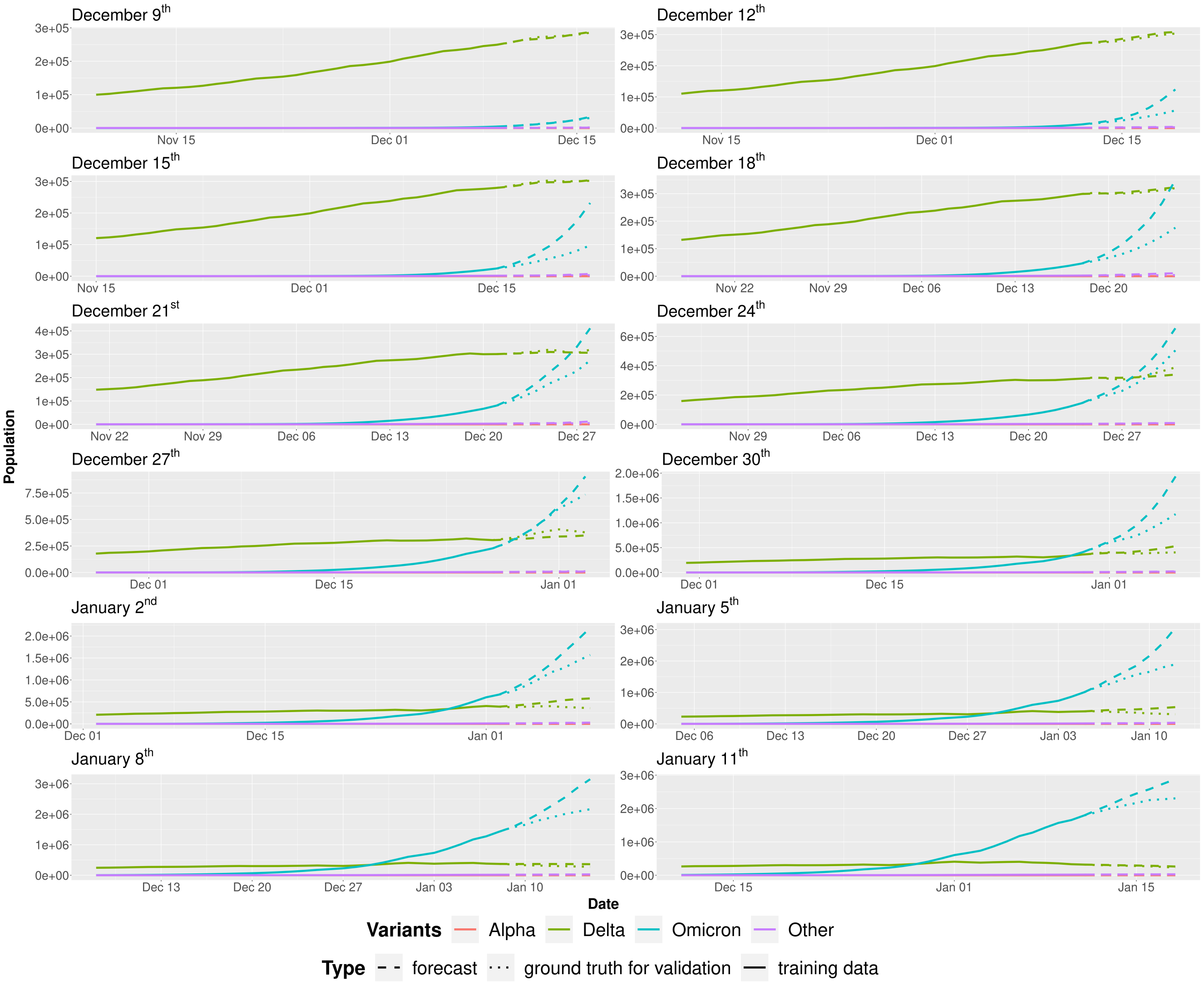


**Figure SM36.** Evolution of the $I_v(\tilde{t})$ compartments using Sybil in the first scenario starting the forecast from December 9th, 12th, 15th, 18th, 21st, 24th, 27th, 30th, 2021 and from January 2nd, 5th, 8th and 11th, 2022 moving the training window by three days (the dashed line shows the prediction, while solid and dotted lines represent the training data and the ground-truth values extracted from the surveillance data, respectively). All plots refer to a forecast one week into the future.

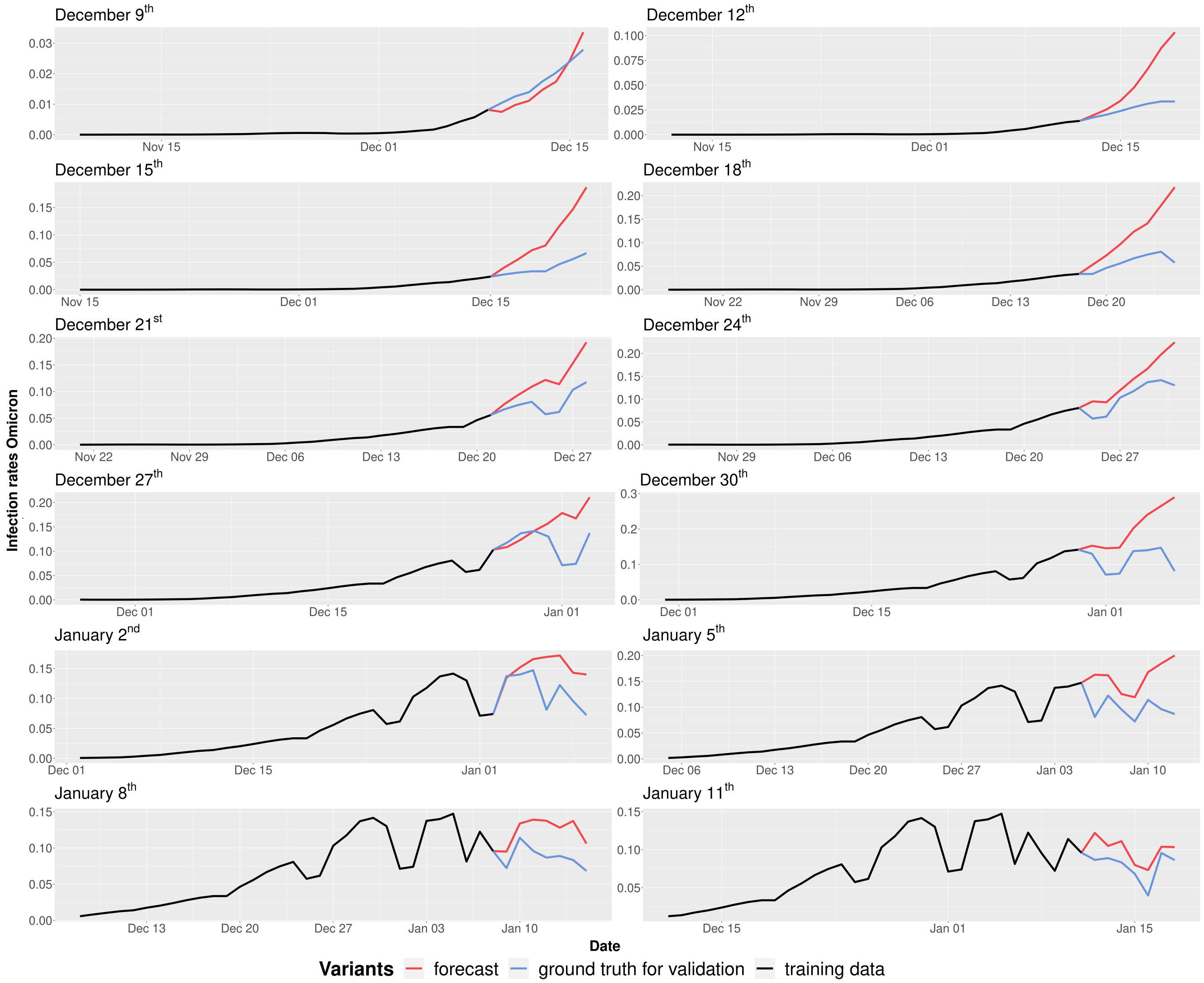


**Figure SM37.** Evolution of the infection rates $\beta_{Omicron}(\tilde{t})$ using Sybil in the first scenario starting the forecast from December 9th, 12th, 15th, 18th, 21st, 24th, 27th, 30th, 2021 and from January 2nd, 5th, 8th and 11th, 2022 moving the training window by three days (the dashed line shows the prediction, while solid and dotted lines represent the training data and the ground-truth values extracted from the surveillance data, respectively). All plots refer to a forecast one week into the future.

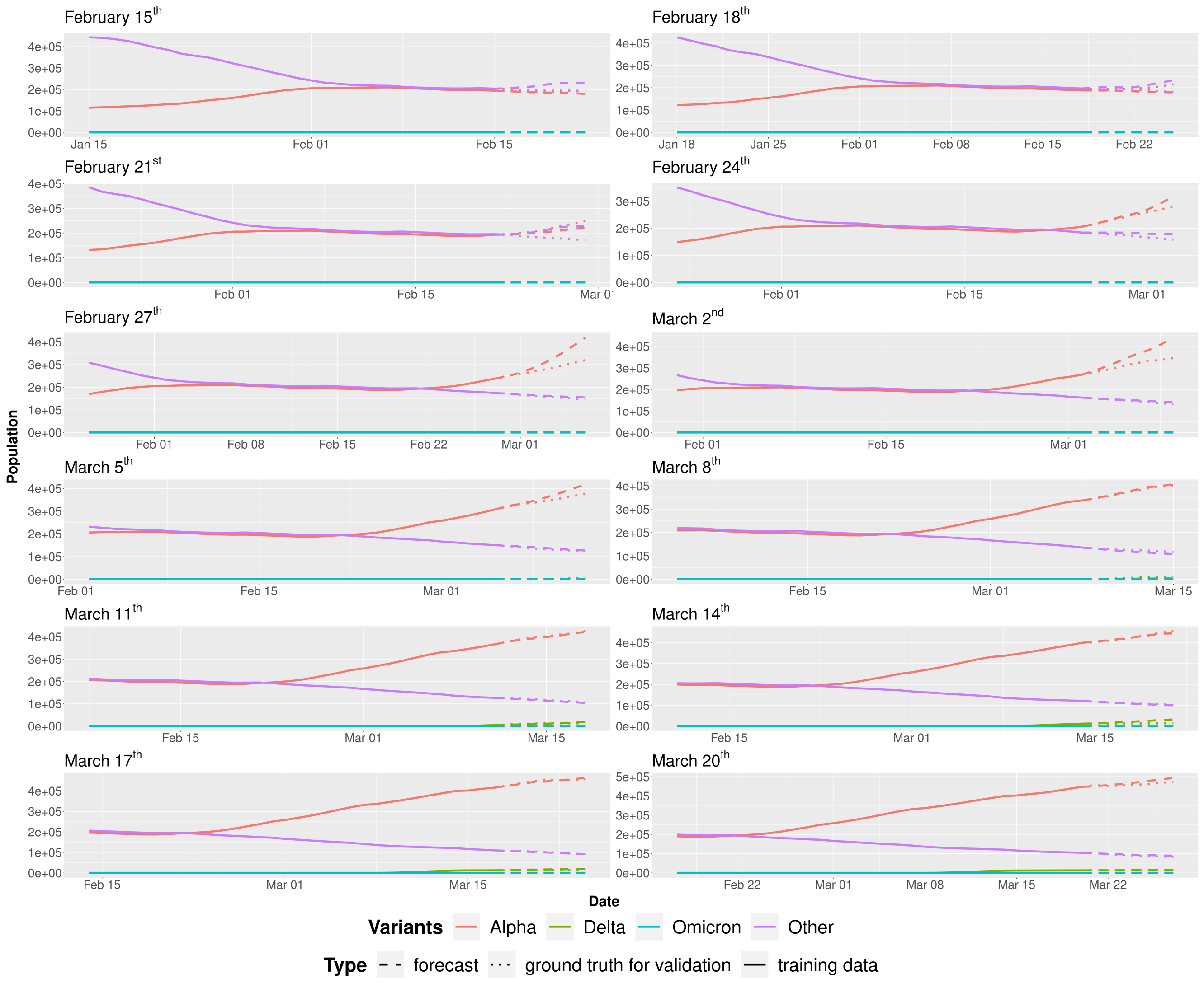


**Figure SM38.** Evolution of the $I_v(\tilde{t})$ compartments using Sybil in the second scenario starting the forecast from February 15$^{th}$, 18$^{th}$, 21$^{st}$, 24$^{th}$, 27$^{th}$, March 2$^{nd}$, 5$^{th}$, 8$^{th}$, 11$^{th}$, 14$^{th}$, 17$^{th}$ and 20$^{th}$, 2021 moving the training window by three days (the dashed line shows the prediction, while solid and dotted lines represent the training data and the ground-truth values extracted from the surveillance data, respectively). All plots refer to a forecast one week into the future.

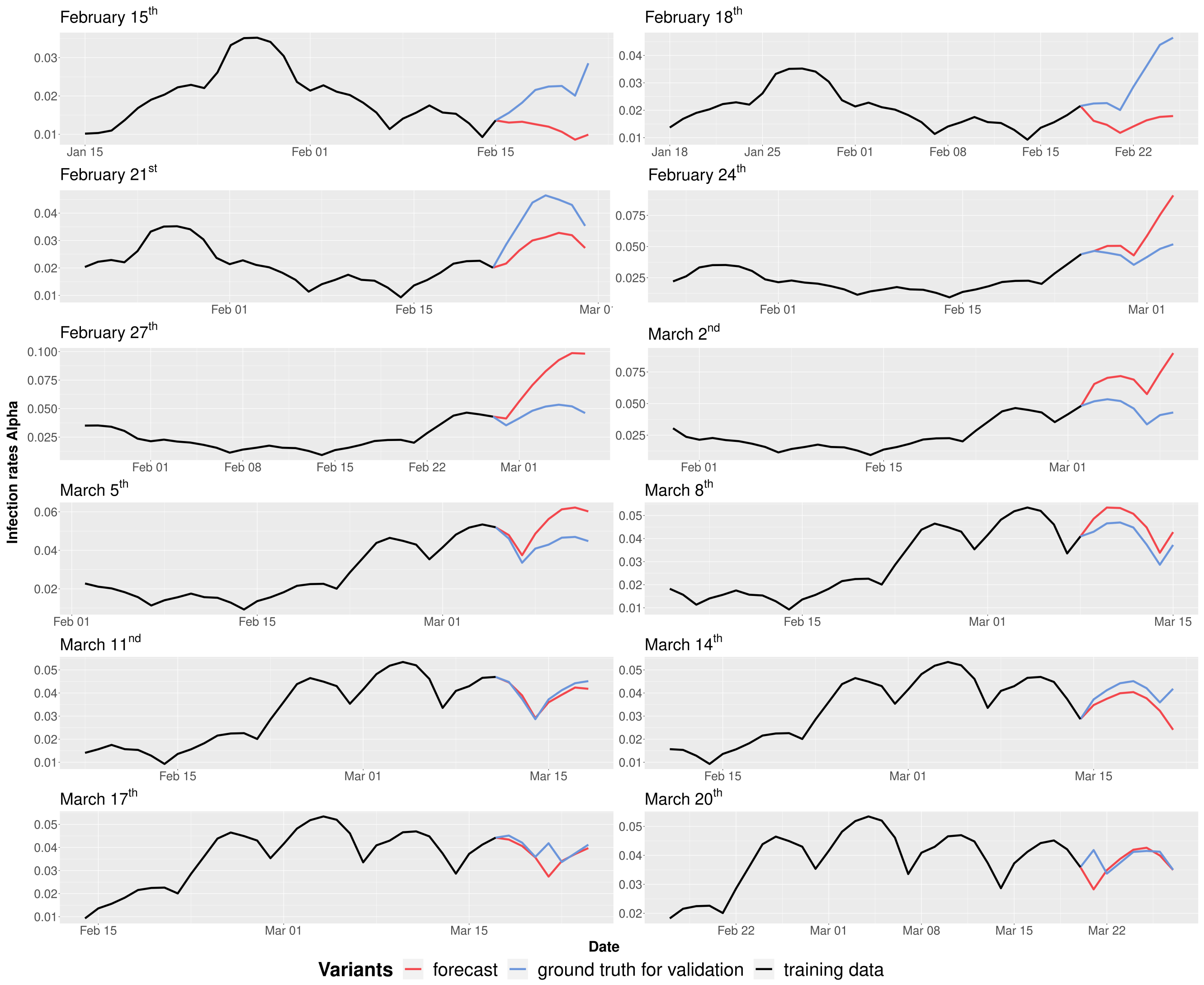


**Figure SM39.** Evolution of the infection rates $\beta_{Alpha}(\tilde{t})$ in the second scenario starting the forecast from February 15$^{th}$, 18$^{th}$, 21$^{st}$, 24$^{th}$, 27$^{th}$, March 2$^{nd}$, 5$^{th}$, 8$^{th}$, 11$^{th}$, 14$^{th}$, 17$^{th}$ and 20$^{th}$, 2021 moving the training window by three days (the dashed line shows the prediction, while solid and dotted lines represent the training data and the ground-truth values extracted from the surveillance data, respectively). All plots refer to a forecast one week into the future.

### 6 How to reproduce the results

#### 6.1 Requirements

You need to have docker installed on your computer, for more info see this document: https://docs.docker.com/engine/installation/. Ensure your user has the rights to run docker (without the use of sudo). To create the docker group and add your user in a Unix system:

- Create the docker group:

```
$ sudo groupadd docker
```

- Add your user to the docker group:

```
$ sudo usermod -aG docker $USER
```

- Log out and log back in so that your group membership is re-evaluated.

##### 6.2 Reproduce

To reproduce the results clone the repository (https://github.com/daniele-baccega/sybil-forecasting.git) and run:

```
$ cd sybil-forecasting
$ ./reproduce.sh
```

To reproduce the results with a fixed recovery rate – see *Fixed recovery rate* subsection – run (inside the sybil-forecasting directory):

```
$ git checkout sybil-v2.0
$ ./reproduce.sh
```
